# Clinical prediction models combining routine clinical measures identify participants with youth-onset diabetes who maintain insulin secretion in the range associated with type 2 diabetes: The SEARCH for Diabetes in Youth Study

**DOI:** 10.1101/2023.09.27.23296128

**Authors:** Angus G Jones, Beverley M Shields, Richard A Oram, Dana M Dabelea, William A Hagopian, Eva Lustigova, Amy S Shah, Julieanne Knupp, Amy K Mottl, Ralph B. D’Agostino, Adrienne Williams, Santica M Marcovina, Catherine Pihoker, Jasmin Divers, Maria J Redondo

## Abstract

**Objective:** With the high prevalence of pediatric obesity and overlapping features between diabetes subtypes, accurately classifying youth-onset diabetes can be challenging. We aimed to develop prediction models that, using characteristics available at diabetes diagnosis, can identify youth who will retain endogenous insulin secretion at levels consistent with type 2 diabetes (T2D).

**Methods:** We studied 2,966 youth with diabetes in the prospective SEARCH study (diagnosis age ≤19 years) to develop prediction models to identify participants with fasting c-peptide ≥250 pmol/L (≥0.75ng/ml) after >3 years (median 74 months) of diabetes duration. Models included clinical measures at baseline visit, at a mean diabetes duration of 11 months (age, BMI, sex, waist circumference, HDL-C), with and without islet autoantibodies (GADA, IA-2A) and a Type 1 Diabetes Genetic Risk Score (T1DGRS).

**Results:** Models using routine clinical measures with or without autoantibodies and T1DGRS were highly accurate in identifying participants with c-peptide ≥0.75 ng/ml (17% of participants; 2.3% and 53% of those with and without positive autoantibodies) (area under receiver operator curve [AUCROC] 0.95-0.98). In internal validation, optimism was very low, with excellent calibration (slope=0.995-0.999). Models retained high performance for predicting retained c-peptide in older youth with obesity (AUCROC 0.88-0.96), and in subgroups defined by self-reported race/ethnicity (AUCROC 0.88-0.97), autoantibody status (AUCROC 0.87-0.96), and clinically diagnosed diabetes types (AUCROC 0.81-0.92).

**Conclusion:** Prediction models combining routine clinical measures at diabetes diagnosis, with or without islet autoantibodies or T1DGRS, can accurately identify youth with diabetes who maintain endogenous insulin secretion in the range associated with type 2 diabetes.

## Introduction

### Accurate identification of diabetes subtype is challenging in youth-onset diabetes

Type 2 diabetes (T2D) is increasingly common in children and young adults, with the prevalence in US youth aged 10 to 19 years more than doubling between 2001 and 2017 Identification of youth-onset T2D can be particularly challenging, as clinical features of type 1 (T1D) and T2D overlap. For example, many children with obesity who develop diabetes have T1D (2). Islet-autoantibody testing may assist classification, especially at diagnosis, but these tests have imperfect specificity and sensitivity, meaning that a positive autoantibody may not confirm T1D, and negative autoantibodies may not exclude this condition (3–5). While c-peptide may assist classification, it is often retained at diagnosis in obese individuals with T1D and in children diagnosed early (e.g., through research study monitoring of autoantibody-positive individuals), limiting utility at diagnosis (6, 7).

### Differences in glycaemic treatment requirements between type 1 and 2 diabetes are largely driven by differences in endogenous insulin secretion

The glycaemic treatment requirements of T1D differ markedly from T2D. These differences are mainly driven by the development of severe insulin deficiency in T1D but not in T2D (6). In T1D, the severe insulin deficiency that occurs in most people living with this condition leads to absolute insulin requirement, need for physiological insulin replacement, high glucose variability and hypoglycaemia risk, and lack of glycaemic benefit from most non-insulin glucose-lowering therapies (6, 8–11). Consistent with this, within individuals with longstanding diabetes assessment of insulin secretion, using c-peptide has been shown to predict successful insulin withdrawal, glucose variability and hypoglycaemia risk, and response to prandial insulin and non-insulin glucose lowering therapies, regardless of clinician diagnosed diabetes subtype (6, 12–18). C-peptide levels over approximately 250pmol/L (0.75ng/ml) fasting or 600pmol/L (1.8ng/ml) stimulated have been suggested to identify individuals with T2D and its treatment requirements, and this threshold has been incorporated into international guidelines (10, 19).

### Prediction models combining clinical features and biomarkers may help identify patients who will retain high c-peptide levels and therefore may be treated as type 2 diabetes

No single feature or biomarker at diagnosis can robustly identify diabetes subtype and future retained endogenous insulin secretion. The interpretation of individual features or biomarker tests such as islet autoantibodies depends on the prior-likelihood, informed by other patient features. For example, islet autoantibodies will have a very low negative predictive value in a child with classical features of T1D, but a high negative predictive value where the clinical features also support an alternative diagnosis. Conversely, a single positive islet autoantibody may have moderate positive predictive value where T1D is unlikely (5, 20).

One way to address this issue is to combine different clinical features and biomarkers using prediction model approaches. In adult-onset diabetes, prediction models with high accuracy have been developed to differentiate T1D and T2D defined by early insulin requirement and longstanding persistence of c-peptide (25,26). The same models have also shown high accuracy for classification of diabetes defined genetically and by pancreatic histology (21, 22). However, no clinical models are available for retained c-peptide or classification in youth-onset diabetes.

We therefore aimed to develop clinical prediction models to combine clinical features and biomarkers, assessed close to diabetes diagnosis, to identify individuals with youth onset diabetes who will retain high levels of endogenous insulin secretion consistent with T2D.

## Methods

We developed prediction models for retained endogenous insulin, consistent with T2D and non-insulin requirement (fasting c-peptide >0.75 ng/mL or 250pmol/L 3 to 10 years after diabetes diagnosis) in participants in the SEARCH for Diabetes in Youth study, a multicentre prospective cohort study (23, 24).

### Cohort

The SEARCH study performed population-based ascertainment of youth with incident diabetes diagnosed under 20 years of age from 2002 through 2020 at five centres in the USA as previously described (23, 24). Participants with recently diagnosed diabetes in 2002–2006 and 2008 participated in a baseline research visit. Those who had a baseline research visit were invited to participate in a follow-up visit (at median diabetes duration 8 years), with serial follow visits undertaken in a subset of participants. At each research visit, fasting blood samples (≥8 hour fast) were obtained. Diabetes type determined by the participant’s healthcare provider was recorded and is referred to as provider type. The SEARCH study was reviewed and approved by the local institutional review boards that had jurisdiction over the local study population, and all participants provided informed consent and/or assent.

For this analysis, participants were included if c-peptide measurement was available between 3 and 10 years (inclusive) post diabetes diagnosis and the following information from the baseline visit was available in the study dataset: age at diagnosis, BMI, waist circumference, sex, HDL-C, GAD and IA2 islet autoantibody status. Those with monogenic diabetes confirmed by genetic testing were excluded from this analysis.

### Model Outcomes

Substantial retained insulin secretion was defined as a fasting c-peptide >250pmol/L (>0.75 ng/mL) between ≥3 and ≤10 years diabetes duration. When c-peptide was <250pmol/L (<0.75ng/mL), concurrent glucose of ≥72mg/dl (4mmol/L) was required for inclusion of the result. Where fasting c-peptide was not available, a random non-fasting c-peptide value was used, converted to the equivalent fasting value by division by 2.5 (6).

### Model Predictors

Model predictor variables were pre-specified based on previously reported associations with differentiating T1D and T2D, availability in the study, and lack of need for fasting blood collection. Models included: sex, BMI, waist circumference, age of diabetes diagnosis, HDL-cholesterol, race/ethnicity, islet-autoantibodies (GADA, IA-2A), and a 67-SNP genetic risk score for genetic susceptibility to type 1 diabetes (T1DGRS2) (25). A 397 SNP genetic risk score for type 2 diabetes (26), and ZnT8 islet-autoantibodies (both available in only a subset of participants) were also assessed for improvement in prediction over and above the above features. Model predictors were assessed at the baseline visit. Where individual parameters were missing at baseline (<0.01% of values for included participants), the closest available measure to diagnosis from later study visits was used. Waist circumference was assessed using the NHANES method (uppermost lateral border of the ilium). Race and ethnicity were self-reported using 2000 US census questions and defined as 4 groups based on prevalence in the SEARCH Study: Black, White-Hispanic, Non-Hispanic White and other (predominantly Asian and Native American).

To provide greatest flexibility for clinical and research use, models were developed and tested with and without variables that are not routinely measured (such as islet autoantibodies and genetic risk scores).

### Laboratory analysis

Diabetes autoantibodies to GAD65 (GADA), IA-2 antigen (IA-2A), and zinc transporter 8 (ZnT8A)), HDL-cholesterol (HDL-C), HbA1c, fasting c-peptide, and glucose were assessed on fasting blood samples at the Northwest Lipid Metabolism and Diabetes Research Laboratories, University of Washington, the central laboratory for SEARCH. The cut-off values for islet-autoantibody positivity were 33 NIDDK units (NIDDKU)/mL for GADA, 5 NIDDKU/mL for IA-2A, and 0.02 NIDDKU/mL for ZnT8A. Serum c-peptide concentration was determined by a two-site immunoenzymetric assay (Tosoh 1800; Tosoh Bioscience).

### Assessment of genetic risk scores

Genotyping and assessment of genetic risk scores in SEARCH have been recently described (26). We calculated two previously published T1DGRS including a 30-SNP score used in many studies (T1DGRS-1(27)) and a more recent 67-SNP score with a greater diversity of HLA variation, including 18 interactions between major HLA class II alleles and additional independent variants (T1DGRS-2(25)). We assessed the T1DGRS-2 in primary analysis. We also assessed a type 2 diabetes GRS (T2DGRS) generated from summary statistics of a previously published comprehensive genome-wide association study in individuals of European ancestry (26, 28).

### Statistical analysis

#### Sample size

We determined sample size using the approach described Riley et al (29) and the pmsampsize package in R (30). For an outcome prevalence of 15%, eight model parameters, and assuming a model optimism-adjusted C-statistic of >0.8, the minimum sample size required for small overfitting (<10% expected shrinkage of predictor effects), small mean average prediction error and small margin of error around estimation of the outcome probability (+/-0.05) was 438. The SEARCH cohort contains 1918 for complete case analysis with all predictors, and thus, well above the minimum required sample size.

#### Model development

Analysis was carried out in STATA version 16.0 (StataCorp, College Station, TX, USA), and R studio version 1.4.1106 (RStudio, PBC, Boston, MA, USA).

Models were developed using logistic regression, on a complete case basis.

We assessed the linearity of the relationship between continuous model variables and outcome on the logit scale using gamplot in R, with and without adjustment for covariates. As all relationships were linear with inclusion of covariates model covariates were not transformed. GADA, IA-2A (and Znt8A) were coded as number of positive islet autoantibodies.

Models were built in the following stages in primary analysis:

1. Clinical characteristics alone (age of diagnosis, BMI, sex, HDL, waist circumference)
2. Clinical characteristics + islet autoantibody number (GADA, IA2)
3. Clinical characteristics + islet autoantibody number + T1DGRS
4. Clinical characteristics + T1DGRS

To maximise utility for both clinical and research use, additional models were developed and reported in supplementary materials with and without waist circumference, HDL-C and ZnT8 islet autoantibody. A T2DGRS was also assessed. Models were developed initially without race/ethnicity to allow generalisability, but final models were tested in separate race/ethnicity subgroups to check for differences in performance. We assessed whether each of race/ethnicity, Znt8 autoantibodies and a T2DGRS improved model performance using the likelihood ratio test, Akaike’s information criterion and Bayesian information criterion.

#### Evaluation of model performance

We assessed model discrimination using AUCROC analysis and precision recall curves and assessed model calibration visually with calibration plots using the STATA module PMCALPLOT (Ensore, Snell, Martin 2018). We also reported model sensitivity, specificity, positive and negative predictive values, and overall classification accuracy for an illustrative binary model cutoff of 50% probability (≥50% assumed retained c-peptide, <50% assumed low c-peptide) and compared this to accuracy of islet autoantibody status alone and latest provider type, with overall classification accuracy compared using the Chi-squared test.

We then assessed model discrimination and calibration for all models separately by race/ethnicity (grouped into Black, white-Hispanic, Non-Hispanic White as above). To determine model performance in situations where clinical diagnosis is challenging, and assess whether models had utility over and above clinical diagnosis and autoantibody testing, we tested models in the following participant subgroups:

1. Older youth (age of onset ≥age 10) with obesity (BMI Z score >1.64)
2. Participants with negative and positive islet-autoantibodies (GADA and IA-2A)
3. Participants with a provider diagnosis of T1D and T2D
4. Within the previously proposed four etiological subtypes of diabetes based on islet antibody status (GADA or IA2A positive or both negative) and presence/absence of insulin resistance (assessed using a formula based on HbA1c, triglycerides and waist circumference, with insulin resistance defined as insulin sensitivity below the 25^th^ centile of 2860 youth without diabetes, as previously reported) (26, 31).

#### Internal validation

Model internal validation was undertaken using bootstrapping (with replacement method, 500 bootstraps) using the RMS (Regression Modelling Strategies) package for R version 6.3 (Frank R Harrell Jr).

## Results

### Participant Characteristics

2,991 participants in SEARCH had measured c-peptide between 3-10 years diabetes duration, of whom 2,966 met analysis inclusion criteria with available data for the routine clinical features model (Model 1) and clinical characteristics with islet autoantibodies model (Model 2). These features and T1DGRS were available in 1918 participants (Models 3 and 4). A flow diagram is shown in Supplementary Figure 1. Overall characteristics of participants included in each model are shown in Supplementary Table 1. For participants included in Model 1 and 2, median (interquartile range (IQR)) diabetes duration at c-peptide assessment was 74 (62–94) months; median (IQR) duration at assessment of model parameters (including islet autoantibodies) was 11(5–18) months. For participants included in Models 1 and 2, race and ethnicity was 65% non-Hispanic White, 14% Hispanic White, 16% Black, and 4% other non-White. The characteristics of participants excluded from models due to missing T1DGRS are shown in Supplementary Table 2. Participants with T1DGRS available, in comparison to those in model 1 and 2 without this measure, were more likely to report Non-Hispanic White ethnicity (69 vs 63%) and less likely to retain c-peptide (20 vs 15%), be islet-autoantibody positive (73 vs 68%) or report type 2 diabetes (18 vs 14%) (p<0.02 for all).

#### Participants with low and retained c-peptide have the characteristics, respectively, of type 1 and 2 diabetes

Characteristics of participants included in models without T1DGRS, by c-peptide outcome status, are shown in Table 1. Retained c-peptide ≥250pmol/L (≥0.75ng/mL) was observed in 17% of participants. Those with retained c-peptide had characteristics largely consistent with type 2 diabetes. T1DGRS and T2DGRS were similar in those with low and high retained c-peptide to values in those with diabetes type defined by a combination of latest provider classification and islet autoantibody testing (Table 1). Fasting c-peptide <250pmol/L (<0.75ng/ml) was highly specific for insulin treatment (specificity >99.9%). In contrast, 47.4% of those with c-peptide ≥250pmol/L were treated without insulin.

**Table 1:** Participant characteristics by C-peptide status. * t-test (continuous variables) or Chi2 test (categorical variables) **BMI Z-score >1.64 indicates obesity. ***NHANES technique, measured at the uppermost lateral border of the ilium.**** Mean T1DGRS2 in participants with autoantibody positive clinically diagnosed T1D and autoantibody negative clinically diagnosed T2D 13.1 and 9.2 respectively. ***** Mean T2DGRS in participants with autoantibody positive clinically diagnosed T1D and autoantibody negative clinically diagnosed T2D 11.1 and 11.4 respectively. ****** Earliest recorded after 3 years diabetes duration, median duration 67 months.

|  | C-peptide <250pmol/L<br>(<0.75ng/ml)<br>(n=2465; 83%) Mean<br>or % (95% CI) | C-peptide ≥250pmol/L<br>(≥0.75ng/ml)<br>(n=501; 17%) Mean or<br>% (95% CI) | P value for<br>comparison* |
| --- | --- | --- | --- |
| Age of onset (years) | 9.5 (9.3, 9.6) | 14.1 (13.9, 13.4) | <0.001 |
| BMI Z-score** | 0.56 (0.52, 0.60) | 2.0 (1.9, 2.1) | <0.001 |
| Waist circumference (cm)*** | 69.2 (68.6, 69.7) | 109.0 (107.0, 111.2) | <0.001 |
| % female | 48.1 (46.1, 50.1)% | 66.9 (62.5, 71.0)% | <0.001 |
| Race and ethnicity | Black 10.9% (n=269)<br>Hispanic-White 12.3% (n=304)<br>Non-Hispanic White 74.3% (n=1830)<br>Other non-white 2.5% (n=61) | Black 43.5% (n=219)<br>Hispanic-White 23.6% (n=118)<br>Non-Hispanic White 21.4% (n=107)<br>Other non-white 11.4% (n=57) | <0.001 |
| Triglycerides (mg/dl) | 65.6 (63.8, 67.6) | 135.6 (124.1, 147.1) | <0.001 |
| HDL (mg/dl) | 55.9 (55.3, 56.4) | 41.9 (41.0, 42.9) | <0.001 |
| GADA or IA2A positive | 83.7% (Both GADA and IA2A positive 42.6%) | 9.8% (both GADA and IA2A positive 3.0%) | <0.001 |
| T1DGRS -2**** | 13.1 (13.0, 13.2) | 9.5 (9.3, 9.7) | <0.001 |
| T2D GRS ***** | 11.1 (11.1, 11.1) | 11.4 (11.3, 11.4) | <0.001 |
| Insulin-treated after 3 years diabetes duration***** | 99.9% | 47.4% | <0.001 |
| Provider type (Clinical classification) | T1D: 97.2%, T2D: 2.3%, other/unknown 0.5% | T1D: 20.4%, T2D: 78.4%, other/unknown 1.6% | <0.001 |

#### Combining features using a diagnostic model improves discrimination over clinical diagnosis or autoantibodies alone

Table 2 shows discrimination performance of models with clinical characteristics alone (age at diagnosis, BMI, waist circumference, sex, HDL-C) (model 1), clinical characteristics with GADA and IA-2A (model 2) and clinical characteristics and T1DGRS with and without islet autoantibodies (models 3 and 4 respectively). Performance of islet autoantibodies alone, and provider diagnosis is also shown for comparison. Model coefficients are shown in Supplementary Table S3.

**Table 2:** Model discrimination and accuracy in identifying participants with retained c-peptide >250pmol/L, in comparison to provider diagnosis and islet autoantibodies alone. Analysis for healthcare provider diagnosis and islet-autoantibody status limited to those with availability of model 1 clinical features. *Using (for retained c-peptide) a ≥50% model probability cutoff, provider diagnosis of 2 diabetes, or absence of islet-autoantibodies. Sensitivity and specificity are for retained c-peptide. ^+^ provider type (latest available) with those a provider diagnosis other than Type 1 or 2 diabetes (n=4) or unknown (n=16) excluded. ^++^AUCROC assessed using number of positive islet-autoantibodies, predictive value and accuracy assessed using any positive islet-autoantibody verses all negative. Note that for islet autoantibodies AUCROC, sensitivity, specificity and predictive values relate to inverted outcome (lower antibody number (AUCROC) or negative islet-autoantibody status (other measures) indicating retained c-peptide.

| Features | AUCROC (95% CI) | Sensitivity* | Specificity* | Predictive value high c-peptide* | Predictive value low c-peptide* | Overall accuracy* (95% CI) |
| --- | --- | --- | --- | --- | --- | --- |
| Routine measures (Model 1) n=2966 (c-peptide >250pmol/L n=501) | 0.950 (0.938, 0.963) | 71.1 (66.9, 75.0)% | 97.4 (96.7, 98.0)% | 85.0 (81.2, 88.2)% | 93.3 (92.2, 94.2)% | 93.0 (92.0, 93.9)% |
| Routine measures & islet-autoantibodies (Model 2) n=2966 (c-peptide >250pmol/L n=501) | 0.972 (0.963, 0.980) | 83.6(80.1, 86.8)% | 98.0 (97.3, 98.5)% | 89.3 (86.2, 92.0)% | 96.7 (95.9, 97.4)% | 95.5 (94.7, 96.3)% |
| Routine Measures & islet-autoantibodies and T1DGRS (Model 3) n=1918 (c-peptide >250pmol/L n=292) | 0.979 (0.969, 0.987) | 86.3 (81.8, 90.0)% | 98.3 (97.6, 98.9)% | 90.3 (86.2, 93.5)% | 97.5 (96.7, 98.2)% | 96.5 (95.6, 97.3)% |
| Routine measures & T1DGRS (Model 4) N=1918 (c-peptide >250pmol/L n=292) | 0.964 (0.952, 0.9760) | 77.1 (71.8, 81.8)% | 97.5 (96.6, 98.2)% | 84.6 (80.1, 88.2)% | 96.0 (95.0, 96.7)% | 94.4 (93.3, 95.4)% |
| Healthcare provider diagnosis+ n=2946 (c-peptide >250pmol/L n=493) | 0.887 (0.869, 0.905) | 79.7 (75.9, 83.2)% | 97.7 (97.0, 98.2)% | 87.3 (83.9, 90.3)% | 96.0 (95.1, 96.7)% | 94.6 (93.7, 95.4)% |
| Islet-autoantibody number/status++ n=2966 (c-peptide >250pmol/L n=501) | 0.878 (0.863, 0.892) | 90.2 (87.3, 92.7)% | 83.7(82.2, 85.2)% | 53.0% (49.6, 56.4)% | 97.7% (96.9, 98.3)% | 84.8% (83.5, 86.1)% |

Models combining routine clinical features, with or without islet-autoantibody testing (GADA, IA-2A) and T1DGRS, were highly discriminative for retained endogenous insulin secretion (Table 2). AUCROC was 0.95 for clinical characteristics alone (Model 1), rising to 0.97 with the inclusion of islet-autoantibodies (Model 2). AUCROC remained similar with the addition of T1DGRS (0.98, Model 3). A model combining clinical features and T1DGRS (without autoantibodies) (Model 4) also showed good discrimination (AUCROC 0.96). In those with T1DGRS available model fit improved with addition of T1DGRS (Likelihood ratio test p <0.0001, Supplementary Table S4).

All models showed good discrimination (Figure 1) and there were few participants with intermediate probabilities in models including islet-autoantibodies or T1DGRS. Calibration was good across all models (Figure 1). Precision recall curves for each model are shown in Supplementary Figure S2; area under the precision recall curve ranged from 0.83 (model 1) to 0.92 (Model 3).

**Figure 1:**
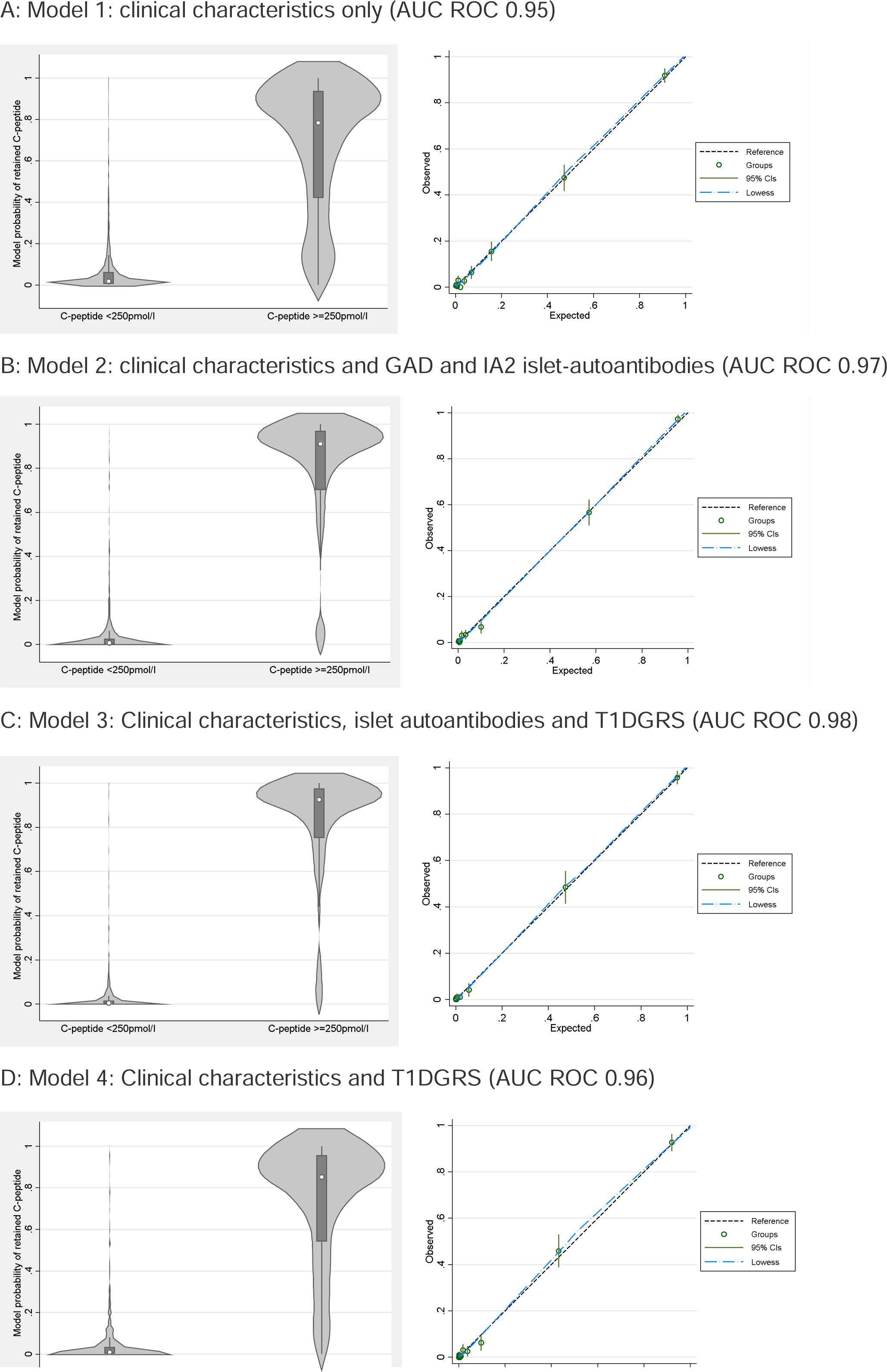
Model separation and calibration. Left column model probability by c-peptide outcome. Right Column model calibration (development dataset): deciles of model predicted probabilities of retained c-peptide plotted against observed c-peptide outcome.

#### All models have substantially better accuracy than islet autoantibody testing alone

All models had higher overall discrimination (AUCROC) than provider type or autoantibodies alone (Table 2 and (for participants with complete data for all models) Supplementary Table S5). Using an arbitrary cut off of 50% probability, all models (including using clinical characteristics only) retained substantially higher overall accuracy than islet autoantibodies alone (p<0.0001 for all).

### Internal validation shows low levels of optimism

All models had similar performance in internal validation, with very low levels of optimism (optimism <0.003 for AUCROC, and <0.033 for calibration slope), suggesting little error due to overfitting (Supplementary Tables S5-S8).

### Models maintain high performance across different racial/ethnic groups

Prevalence of retained c-peptide varied markedly by race/ethnicity: 45%, 28% and 6% of those reporting black, white Hispanic and non-white Hispanic race/ethnicity retained c-peptide ≥250pmol/L (model 1 and 2 cohort). Discrimination and classification accuracy of models 1-4 in these ethnicities, in comparison to islet-autoantibodies and clinical provider diagnosis alone are shown in Supplementary Table S8. For models without T1DGRS, discrimination performance was lower in Non-Hispanic White ethnicity participants (model 1 and 2 AUCROC 0.88 and 0.94 respectively) in contrast to Black (AUCROC 0.95 and 0.97) and white-Hispanic (AUCROC 0.94 and 0.97) participants. Across all ethnicities AUCROC and accuracy using a single (50% probability) cut off was substantially higher for models incorporating islet autoantibodies than clinical diagnosis or autoantibody status alone.

Model calibration by race and ethnicity (Supplementary Figure S3) shows that Model 1 (routine clinical features) moderately underestimated probability of retained insulin secretion for Black participants with intermediate probability but had good calibration for most participants with very high or low probability. In contrast, in non-Hispanic White participants, probability of retained endogenous insulin secretion was moderately overestimated, consistent with the very low prevalence of retained c-peptide (and reported T2D) in this group. Calibration was good across all ethnicities in models incorporating islet antibodies and/or T1DGRS (Models 2-4), with the exception of modest overestimation of retained c-peptide for model 2 in those reporting Non-Hispanic White ethnicity (Figure 2 (model 2) and Supplementary Figure S3 (models 1, 3, and 4)).

**Figure 2:**
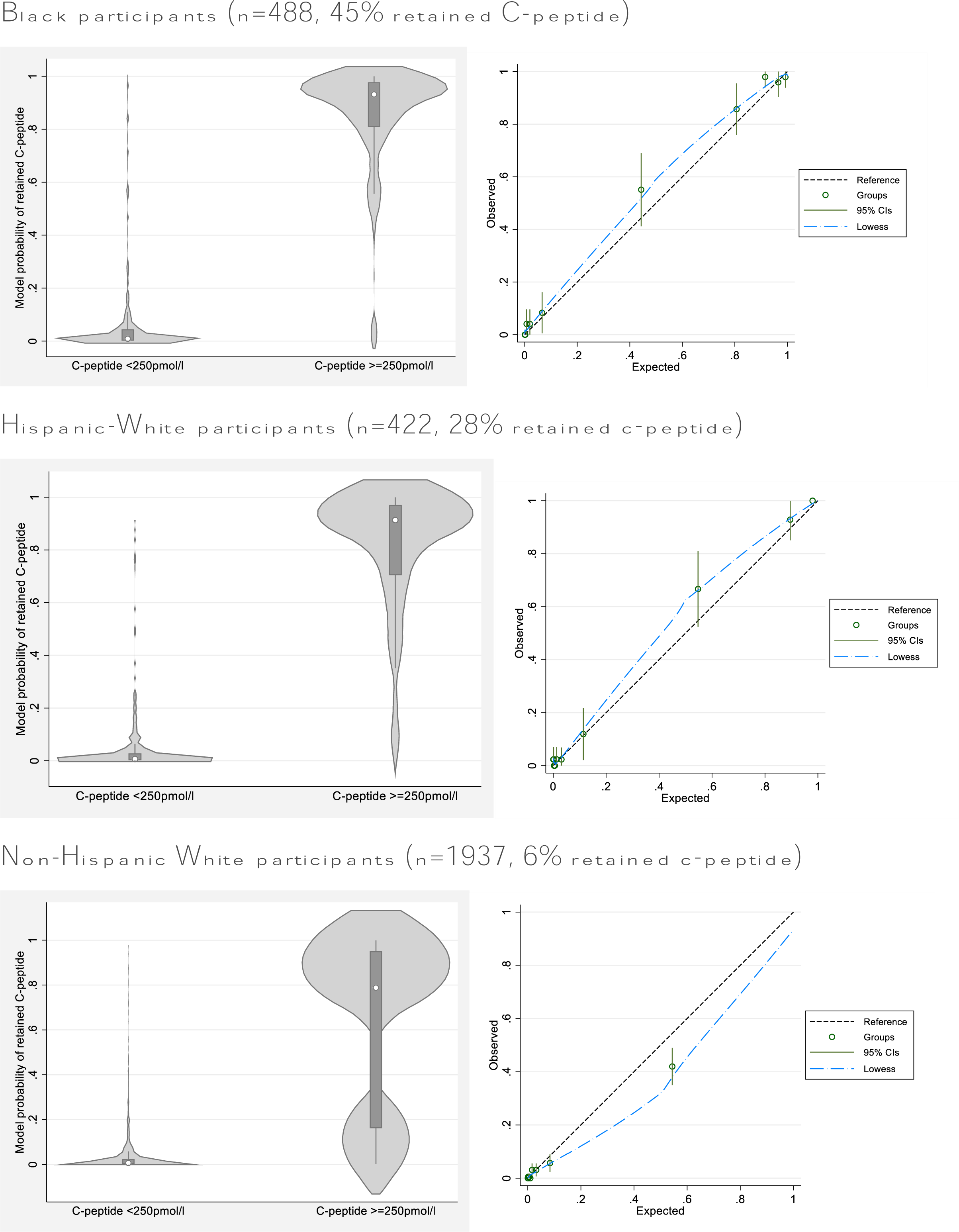
Model 2 (clinical characteristics and islet autoantibodies) separation and calibration by race/ethnicity. Left column model probability by c-peptide outcome. Right column model calibration (development dataset): deciles of model predicted probabilities of retained c-peptide plotted against observed c-peptide outcome.

Performance of models incorporating race/ethnicity are shown in Supplementary Table S10 and Supplementary Figure 4. While incorporating race and ethnicity modestly improved model fit (Likelihood ratio test p<0.04 for all models) adding race and ethnicity did not meaningfully change overall discrimination.

### Models retain high performance in older children with obesity

In youth with age at diagnosis ≥10 years and obesity (BMI Z score >1.64) (a challenging group to classify), models retained high performance with AUCROC 0.86, 0.95, 0.96 and 0.91 for Models 1-4 respectively, and high calibration (Supplementary Table S10 and Supplementary Figure S6). In this group (of whom 70% retained c-peptide), overall accuracy of provider type for retained c-peptide was modest (87%) and superseded by islet-autoantibodies alone (accuracy 90.1%), and (using an illustrative 50% probability cut-off) models incorporating islet-autoantibody testing (Model 2 and 3 accuracy 92.1% and 92.2% respectively) (p vs provider diagnosis <0.05 for all).

### Models are highly predictive of retained c-peptide within those with negative and positive islet-autoantibodies

In participants with negative autoantibodies (53% (452/853) of who retained c-peptide) model 2 and 3 maintained high discrimination (AUCROC 0.95 (95% CI 0.94, 0.97) and 0.96 (0.95, 0.98) respectively) and good calibration (Supplementary Figure S7). Within those with positive islet-autoantibodies, models 2 and 3 also maintained good discrimination (AUCROC 0.86 (0.80, 0.92) and 0.87 (0.78, 0.950) and reasonable calibration (Supplementary Figure S7). However only 49 of 2213 included islet-autoantibody positive participants, retained C-peptide (3.2% and 1.4% of those with 1 and 2 positive islet autoantibodies respectively).

### Models maintain utility for identifying those who will retain c-peptide within clinically and aetiologically defined diabetes subtypes

Within those with clinically diagnosed T1D and T2D (latest available provider type) models retained the ability to discriminate those with and without retained c-peptide. In those diagnosed as T1D (of whom 100 of 2496 (Model 1 and 2) and 57 of 1640 (Model 3 and 4) retained c-peptide) AUCROC (95% CI) was 0.84 (0.78, 0.89), 0.90 (0.86, 0.93), 0.92 (0.88, 0.97) and 0.90 (0.86, 0.94) for models 1-4 respectively. In those diagnosed as T2D (393 of 450 (Model 1 and 2) and 228 of 263 (Model 3 and 4) retained c-peptide) AUCROC was 0.81 (0.76-0.87), 0.87 (0.82-0.93), 0.877 (0.78, 0.95) and 0.82 (0.72, 0.91).

Within those with four subtypes of diabetes defined by the SEARCH classification system (based on autoantibody status and insulin resistance) models also maintained high discrimination for retained c-peptide >250pmol/L: AUCROC ranged from 0.82-0.86 in those islet-autoantibody positive and insulin sensitive, 0.87-0.93 those islet-autoantibody positive and insulin resistant, 0.86-0.91 in those islet-autoantibody negative and insulin sensitive, and 0.86-0.90 in those islet-autoantibody negative and insulin resistant (Supplementary Table S11).

### Impact of ZnT8 autoantibody testing and type 2 diabetes genetic risk score

The addition of ZnT8 autoantibodies (available in 2767 and 1888 included participants for model 1-2 and 3-4 respectively) improved model fit (likelihood ratio test p<0.0001) but did not meaningfully change overall discrimination (Supplementary Table S12). The addition of a type 2 diabetes genetic risk score marginally improved model fit (likelihood ratio test p value 0.03 (model 3) and <0.0001 (model 4)) with no impact on overall discrimination (Supplementary Table S13). Models without waist circumference and HDL maintained high performance (AUCROC 0.94-98): Supplementary table S14. Use of a 30 SNP T1DGRS in place of a 67 SNP score did not meaningfully change performance of models incorporating T1DGRS, with essentially identical model discrimination: Supplementary Table S15.

### Online calculator and model coefficients

An online calculator (beta version) is available on https://julieanneknupp.shinyapps.io/AngusSEARCH/. Model coefficients for all models are shown in Supplementary Table S16.

## Discussion

We have developed and evaluated prediction models to estimate the probability of a patient with youth-onset diabetes maintaining long term c-peptide at levels associated type 2 diabetes and glycaemic treatment requirements. These models show high performance, outperforming islet autoantibodies or clinical diagnosis alone, and could potentially assist classification and treatment decisions in clinical practice. Importantly models retained high performance in hard to classify groups, within those with type 1 or 2 diabetes defined by clinical diagnosis or aetiological markers, and within those of different race/ethnicity.

To our knowledge this is the first study to develop a prediction model for retained c-peptide or diabetes classification in children and adolescents. Our findings are consistent with the high performance of models developed in adult-onset diabetes for classifying diabetes subtype based on a combination of early insulin treatment and retained c-peptide, or pancreatic histology, in a European/American population, and on a combination of islet-autoantibodies and c-peptide in a Chinese population (21, 22, 32–34). These previous models were developed on cross sectional data, and the available data close to diagnosis in SEARCH allows prospective prediction of retained c-peptide, a key advantage in comparison to previous work. The data available in SEARCH also allowed inclusion of a wider range of features.

The robust performance of models including T1DGRS in those of different ethnicities, and improvement in calibration with this marker, provide additional evidence for use of this score in multiple racial and ethnic groups, and is consistent with recent work demonstrating utility across ethnicities in a US population (35). However, the gains seen with a T1DGRS are more modest than previously reported, likely because in SEARCH, the combination of available clinical features and islet-autoantibodies close to diagnosis provide very high accuracy, with limited room for further improvement in discrimination measures like AUCROC. As previously demonstrated, the clinical utility of these risk scores is greatest in those who are hard to classify on other features, for example islet-autoantibody negative suspected T1D, or those with single positive antibodies and suspected T2D (36, 37).

Strengths of this research include a large sample size and availability of prospective data. The large sample size available in SEARCH, combined with the modest number of pre-defined predictors included, means that the risk of overfitting (a common problem in prediction model development) is low, as evidenced by very similar model performance in internal validation. Additional strengths include diversity in race and ethnicity, allowing assessment of model performance across three race and ethnicity groups.

A key limitation is the lack of other datasets for external validation. While robust internal validation suggests high performance without overfitting, further validation assessing model performance in different settings is an important area of further research. In particular, assessment of performance in other race/ethnicity groups is needed. For example, our findings of high discrimination and calibration across different race and ethnicities may not apply to those of South Asian or Native American heritage, who have a particularly high risk of young onset T2D. Lastly, the T1DGRS used in this study was developed in a White-European population, and it is possible that ancestry specific scores may further improve performance (38).

The outcome of these models was based on a c-peptide threshold that is associated with discriminating T2D from T1D and identifying lack of insulin requirement in previous insulin withdrawal studies. These thresholds are derived predominantly from adult studies, and there are limited data validating this threshold in children: a modestly higher threshold has been proposed for identifying T2D at diagnosis in a large Swedish study (39), but data in longstanding diabetes are lacking. The extremely high specificity of this threshold for insulin requirement in SEARCH gives some confidence that children who develop c-peptide below our cut off level will require insulin. However it is possible that a higher threshold may be appropriate for identifying patients who benefit from treatment approaches usually used in T2D in children, as youth developing T2D are generally more obese and insulin resistant than adults with this condition (40).

### Implications

These models have the potential to assist clinicians to determine whether a patient with recently diagnosed youth-onset diabetes is likely to maintain insulin secretion, and therefore can be treated as type 2 diabetes (potentially without insulin). Clinical features only models may also help determine if classification biomarker testing (autoantibodies and/or T1DGRS) is indicated. Importantly, tools of this nature are decision aids to supplement clinical judgement which will incorporate additional information important to decision making, for example glycaemia and patient preference. The provision of continuous probabilities, rather than a binary result, allows clinicians to recognise uncertainty and act accordingly. Optimal use is likely to be through a staged approach at diabetes diagnosis, with classification biomarker testing prioritised in those with intermediate probability. In longstanding diabetes, c-peptide can be measured directly at modest cost and these models may potentially assist in prioritisation of testing.

Models may also be useful for researchers who need to identify diabetes subtypes in research cohorts, and may have particular utility where robust clinical diagnosis is not available (common in electronic healthcare records), and where genetic but not islet-autoantibody data is available, common in many biobanks (22).

In conclusion, prediction models combining routine clinical measures, with or without islet-autoantibodies and a T1DGRS, can accurately identify youth with diabetes who maintain endogenous insulin secretion in the range associated with type 2 diabetes treatment requirements.

## Acknowledgments

The SEARCH study investigators are indebted to the many youth and their families and health care providers whose participation made this study possible.

## Author Contributors

AGJ, BMS, RAO, JD and MJR designed the study. DMD, WAG, EL, AS, AKM, RBD, AW, SMM, CP, JD and RAO researched the underlying data. AGJ analyzed the data with assistance from BMS, JK and JD. AGJ wrote the first draft of the report. All authors provided helpful discussion and reviewed and edited the manuscript. AGJ is the guarantor of this work and, as such, had full access to all the data in the study and takes responsibility for the integrity of the data and the accuracy of the data analysis.

### Prior Presentation

Early data from this study were presented in abstract form at the 82nd Scientific Sessions of the American Diabetes Association, June 2019.

## Data Availability

Data from the SEARCH Study is publcially available though the NIDDK central repository (https://repository.niddk.nih.gov/studies/search/) with additional data (such as genetic risk scores) available through application to the SEARCH study steering committee.

## Ethics statement

The study was reviewed and approved by the local institutional review boards that had jurisdiction over the local study population, and all participants provided informed consent and/or assent.

