## Supplementary for "Clinical prediction models combining routine clinical measures identify participants with youth-onset diabetes who maintain insulin secretion in the range associated with type 2 diabetes: The SEARCH for Diabetes in Youth Study"

**Supplementary Materials**

**Supplementary Figure S1: Study flow diagram**

No available c-peptide measurement within 3-10 years diabetes duration: n = 2567

*(Latest follow up <3 years n = 1880, missing valid c-peptide at potentially eligible visit n = 696)*

Total SEARCH Cohort: **n=5567**

Measured c-peptide between 3- and 10-years diabetes duration: **n=2991**

Missing core clinical features or GAD and IA2 islet-autoantibodies: n = 25

### Included in Model 1 and 2:

**n=2966**

### (Model 1 clinical features only, Model 2 clinical features and islet-autoantibodies)

Meeting criteria for Model 1 and 2 but missing T1DGRS: n=1048

### Included in Models 3 and 4 (incorporating T1DGRS):

**n=1918**

**Supplementary Table S1: Overall participants characteristics for models without T1DGRS (Model 1 and 2, n=2966) and Models with T1DGRS (Model 3 and 4, n=1918).**

*BMI *Z-score >1.64 indicates obesity. **NHANES technique, measured at the uppermost lateral border of the ilium*

|  | Model 1 and 2 (n=2966)  Mean (95% CI) or % | Model 3 and 4 (N=1918)  Mean (95% CI) or % |
| --- | --- | --- |
| c-peptide >250pmol/L (at 3-10 years diabetes duration) | 16.9% | 15.2% |
| Age of onset (years) | 10.3 (10.09, 10.4) | 10.2 (10.0, 10.39) |
| BMI Z-score* | 0.80 (0.76, 0.84) | 0.80 (0.75, 0.85) |
| Waist circumference (cm)** | 75.9 (75.1, 76.7) | 75.4 (74.5, 76.4) |
| % female | 51.3% | 51.0% |
| Race and ethnicity | Black 16.5%  Hispanic 14.2%  White European 65.3%  Other 4.0% | Black 18.3%  Hispanic 16.6%  White European 63.3%  Other 1.8% |
| Triglycerides (mg/dl) | 78.1 (75.3, 80.8) | 75.8 (72.7, 78.8) |
| HDL (mg/dl) | 53.5 (53.0, 54.0) | 53.9 (53.3, 54.5) |
| GADA or IA2A positive | 71.2% (both positive 35.9%) | 73.1% (both positive 37.2%) |
| Clinical classification | Type 1 84.2%,  Type 2 15.2%,  Other 0.7% | Type 1 85.5%,  Type 2 13.7%,  Other 0.8% |

**Supplementary Table S2: Comparison of included participants to those excluded from Models 3 and 4 due to missing T1DGRS.**

*BMI *Z-score >1.64 indicates obesity. **NHANES technique, measured at the uppermost lateral border of the ilium* *** *t-test (continuous variables) or Chi2 test (categorical variables)*

|  | **Missing T1DGRS (Excluded from Models 3 and 4, n=1048)**  Mean (95% CI) or % (95% CI) | **With T1DGRS (Included for Models 3 and 4, n=1918)**  Mean (95% CI) or % (95% CI) | **P value***** |
| --- | --- | --- | --- |
| C-peptide >250pmol/L (at 3-10 years diabetes duration) | 19.9% (17.6, 22.5) | 15.2% (13.6, 16.9) | 0.001 |
| Age of onset (years) | 10.3 (10.1, 10.6) | 10.2 (10.0, 10.39) | 0.4 |
| BMI Z-score* | 0.82 (0.75, 0.89) | 0.80 (0.75, 0.85) | 0.6 |
| Waist circumference (cm)** | 76.8 (75.3, 78.2) | 75.4 (74.5, 76.4) | 0.1 |
| % female | 51.7% | 51.0% | 0.7 |
| Race and ethnicity | Black 13.2%  Hispanic 9.8%  White European 69.1%  Other 7.9% | Black 18.3%  Hispanic 16.6%  White European 63.3%  Other 1.8% | <0.001 |
| Triglycerides (mg/dl) | 82.1 (76.6, 87.7) | 75.8 (72.7, 78.8) | 0.03 |
| HDL (mg/dl) | 52.7 (51.9, 53.5) | 53.9 (53.3, 54.5) | 0.002 |
| GADA or IA2A positive | 67.7% (both positive 33.4%) | 73.1% (both positive 37.2%) | 0.06 |
| Clinical classification | Type 1 81.8%  Type 2 17.8%  Other 0.4% | Type 1 85.5%,  Type 2 13.7%,  Other 0.8% | 0.02 |

**Supplementary Table S3: Models 1 to 4 coefficients and covariate statistical significance.**

**A: Model 1 (n=2966)**

|  | **Beta coefficient** | **95% Confidence interval** | **P value** |
| --- | --- | --- | --- |
| Male sex | -0.8700783 | -1.186169,  -0.5539874 | <0.001 |
| Age at diagnosis (years) | 0.0739423 | 0.0227741, 0.1251105 | 0.005 |
| BMI (kg/m^2^) | 0.1592899 | 0.1092293, 0.2093506 | <0.001 |
| HDL (mg/dl) | -0.0529831 | -0.0676305,  -0.0383357 | <0.001 |
| Waist circumference (cm) | 0.0368719 | 0.0171108, 0.056633 | <0.001 |
| Constant (intercept) | -6.843912 | -8.132948,  -5.554876 | <0.001 |

**B: Model 2 (n=2966)**

|  | **Beta coefficient** | **95% Confidence interval** | **P value** |
| --- | --- | --- | --- |
| Male sex | -0.9339223 | -1.322133,  -0.545712 | <0.001 |
| Age at diagnosis (years) | 0.1345481 | 0.0743977, 0.1946985 | <0.001 |
| BMI (kg/m^2^) | 0.138903 | 0.0822534, 0.1955527 | <0.001 |
| HDL (mg/dl) | -0.0435481 | -0.0606203,  -0.0264759 | <0.001 |
| Waist circumference (cm) | 0.0209935 | -0.0011661, 0.0431531 | 0.06 |
| One positive autoantibody (GADA or IA2-A, verses both negative) | -2.856424 | -3.320372,  -2.392476 | <0.001 |
| Two positive autoantibodies (GAD and IA2-A, verses both negative) | -3.963131 | -4.572985,  -3.353277 | <0.001 |
| Constant (intercept) | -4.502606 | -5.950835,  -3.054377 | <0.001 |

**C: Model 3 (n=1918)**

|  | **Beta coefficient** | **95% Confidence interval** | **P value** |
| --- | --- | --- | --- |
| Male sex | -0.7741532 | -1.31814,  -0.2301662 | 0.005 |
| Age at diagnosis (years) | 0.1542869 | 0.0683962, 0.2401776 | <0.001 |
| BMI (kg/m^2^) | 0.108483 | 0.0378147, 0.1791513 | 0.003 |
| HDL (mg/dl) | -0.0369311 | -0.059891,  -0.0139711 | 0.002 |
| Waist circumference (cm) | 0.0177613 | -0.0097904, 0.045313 | 0.2 |
| One positive autoantibody (GADA or IA2-A, verses both negative) | -2.725245 | -3.362004,  -2.088485 | <0.001 |
| Two positive autoantibodies (GAD and IA2-A, verses both negative) | -3.751959 | -4.620971,  -2.882948 | <0.001 |
| T1DGRS2 | -0.3859143 | -0.5006659,  -0.2711627 | <0.001 |
| Constant (intercept) | 0.1216426 | -2.228487,  2.471772 | 0.9 |

**D: Model 4 (n=1918)**

|  | **Beta coefficient** | **95% Confidence interval** | **P value** |
| --- | --- | --- | --- |
| Male sex | -0.8380161 | -1.295343,  -0.3806892 | <0.001 |
| Age at diagnosis (years) | 0.0896181 | 0.0169049, 0.1623314 | 0.016 |
| BMI (kg/m^2^) | 0.1171035 | 0.0488291, 0.1853779 | 0.001 |
| HDL (mg/dl) | -0.0481782 | -0.0674985,  -0.0288579 | <0.001 |
| Waist circumference (cm) | 0.0292977 | 0.0019118, 0.0566836 | 0.036 |
| T1DGRS2 | -0.5689743 | -0.6706761,  -0.4672725 | 0.000 |
| Constant (intercept) | 0.7560816 | -1.319982,  2.832146 | 0.5 |

**Supplementary Table S4: Change in model performance with inclusion of T1DGRS (over and above Model 2 – clinical features and islet antibodies)**

Likelihood ratio test p<0.0001

| **Model** | **n** | **Akaike's information criterion** | **Bayesian information criterion** |
| --- | --- | --- | --- |
| Routine clinical features and GAD/IA2 (no T1DGRS) | 1918 | 498.5198 | 542.9921 |
| Routine clinical features and GAD/IA2 with T1DGRS | 1918 | 451.648 | 501.6794 |

**Supplementary Table S5: Model discrimination and accuracy in identifying participants with retained c-peptide >250pmol/L, in comparison to provider diagnosis and islet autoantibodies alone, limited to participants with complete data for all models.**

*Using (for retained c-peptide) a ≥50% model probability cut off, provider diagnosis of Type 2 diabetes, or absence of islet-autoantibodies. Sensitivity and specificity are for retained c-peptide.

+ provider type with those a provider diagnosis other than Type 1 or 2 diabetes or unknown excluded

++ AUC ROC assessed using number of positive islet-autoantibodies, predictive value and accuracy assessed using any positive islet-autoantibody verses all negative.

Note that for islet autoantibodies AUC ROC, sensitivity, specificity and predictive values relates to inverted outcome (lower antibody number (AUC ROC) or negative islet-autoantibody status (other measures) indicating retained c-peptide).

| **Features** | **AUC ROC**  **(95% CI)** | **Overall accuracy* (95% CI)** |
| --- | --- | --- |
| Routine measures  (Model 1) n=1918 | 0.943 (0.928, 0.959) | 93.2 (92.0, 94.3) % |
| Routine measures + islet-autoantibodies  (Model 2) n=1918 | 0.975 (0.965, 0.985) | 95.8 (94.8, 96.7) % |
| Routine Measures + islet-autoantibodies and T1DGRS  (Model 3) n=1918 | 0979 (0.969, 0.987) | 96.5 (95.6, 97.3) % |
| Routine measures + T1DGRS  (Model 4)  n=1918 | 0.964 (0.952, 0.9760 | 94.4 (93.3, 95.4) % |
| Healthcare provider diagnosis^+^  n=1902 | 0.889 (0.866, 0.913) | 95.2 (94.1, 96.1) % |
| Islet-autoantibody number or status^++^  n=1918 | 0.888 (0.971, 0.905) | 85.7 (84.0, 87.2) % |

**Supplementary Figure S2: Model 1-4 precision recall curves**. Positive predictive value (‘precision’) for retained endogenous insulin secretion (c-peptide >250pmol/L) plotted against sensitivity (‘recall’) across the range of model probability.

**Model 1: Area under the precision recall curve 0.837**

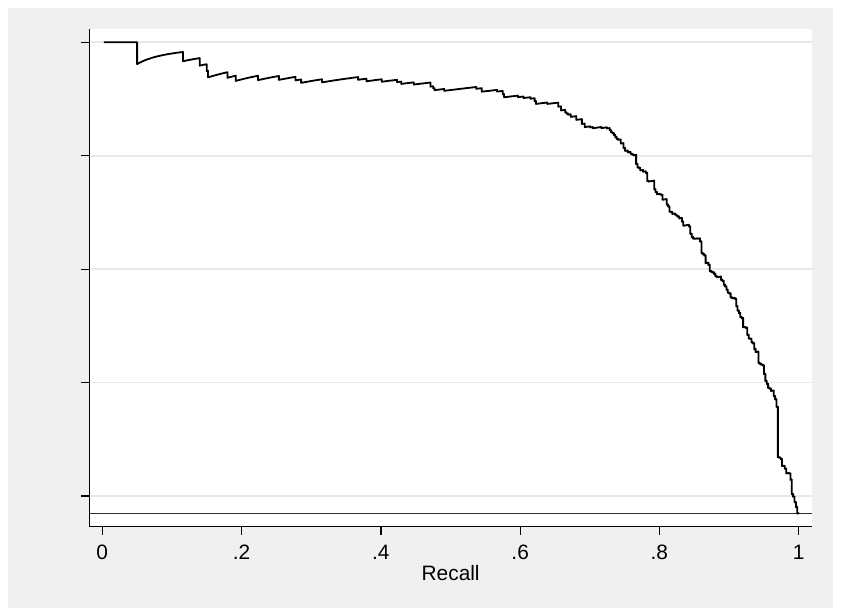

**Model 2: Area under the precision recall curve 0.915**

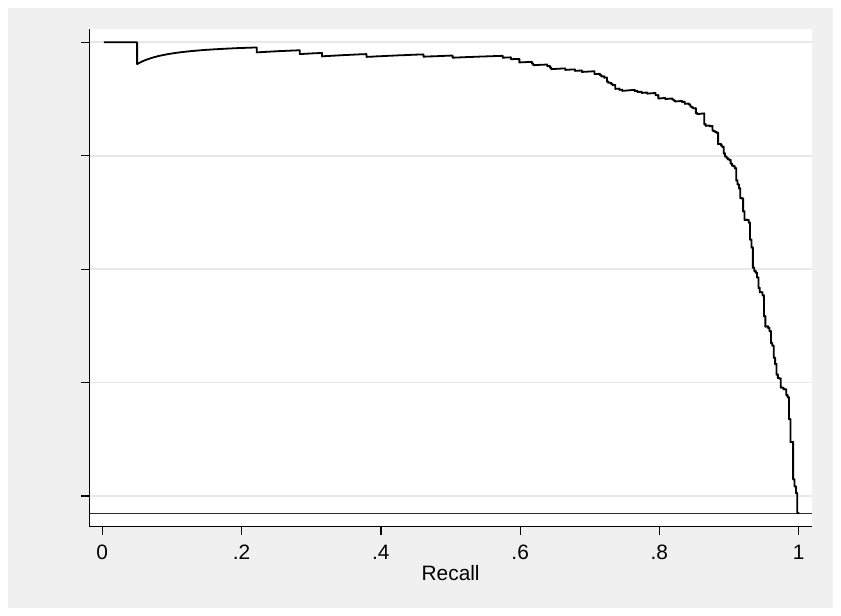

**Model 3: Area under the precision recall curve 0.918**

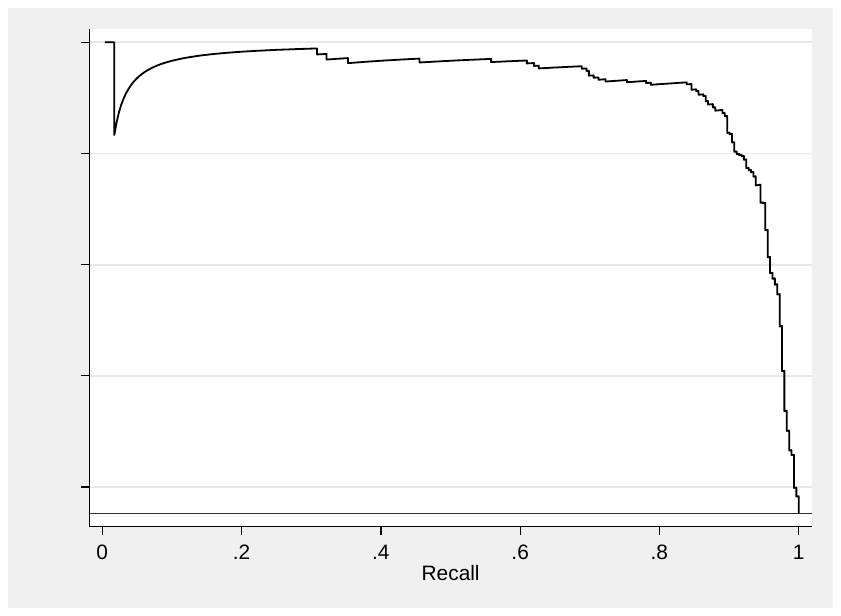

**Model 4: Area under precision recall curve 0.862**

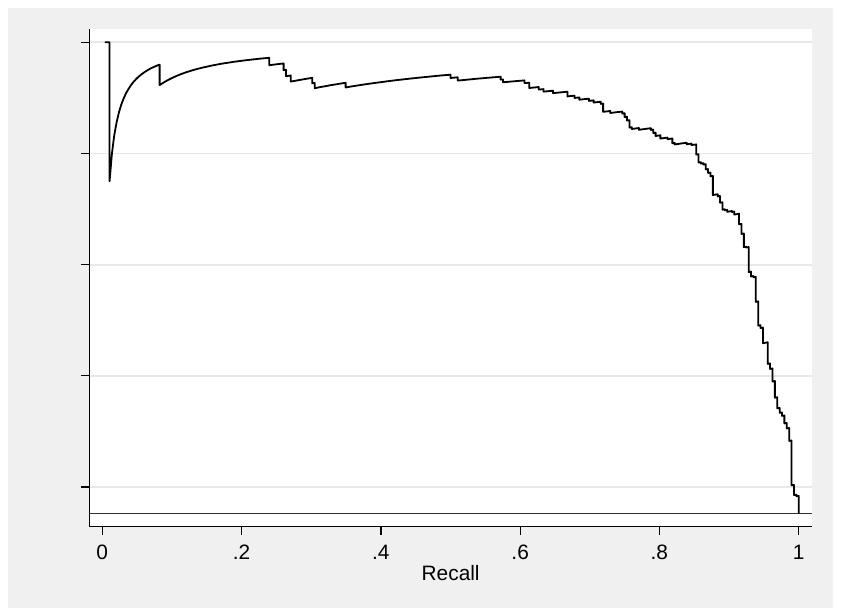

**Supplementary Table S5: Internal validation Model 1 (routine clinical features only):**

|  | **Training** | **Test** | **Optimism** |
| --- | --- | --- | --- |
| **AUC ROC** | 0.9452 | 0.9451 | 0.0001 |
| **Sommers D** | 0.8913 | 0.8902 | 0.0011 |
| **R2** | 0.6691 | 0.6668 | 0.0024 |
| **Intercept** | 0.0000 | 0.0039 | -0.0039 |
| **Slope** | 1.0000 | 0.9944 | 0.0056 |
| **Emax** | 0.0000 | 0.0019 | 0.0019 |

**Supplementary Table S6: Internal validation Model 2 (routine clinical features and GAD and IA2 islet-autoantibodies):**

|  | **Training** | **Test** | **Optimism** |
| --- | --- | --- | --- |
| **AUC ROC** | 0.9743 | 0.9732 | 0.0011 |
| **Sommers D** | 0.9486 | 0.9464 | 0.0022 |
| **R2** | 0.7976 | 0.7931 | 0.0045 |
| **Intercept** | 0.0000 | -0.0093 | 0.0093 |
| **Slope** | 1.0000 | 0.9862 | 0.0138 |
| **Emax** | 0.0000 | 0.0046 | 0.0046 |

**Supplementary Table S7: Internal of validation Model 3 (routine clinical features and GAD, and IA2 islet-autoantibodies and T1DGRS):**

|  | **Training** | **Test** | **Optimism** |
| --- | --- | --- | --- |
| **AUC ROC** | 0.9809 | 0.9785 | 0.0024 |
| **Sommers D** | 0.9617 | 0.9570 | 0.0047 |
| **R2** | 0.8222 | 0.8121 | 0.0101 |
| **Intercept** | 0.0000 | -0.0249 | 0.0249 |
| **Slope** | 1.0000 | 0.9643 | 0.0357 |
| **Emax** | 0.0000 | 0.0123 | 0.0357 |

**Supplementary Table S8: Internal validation of Model 4 (routine clinical features and T1DGRS):**

|  | **Training** | **Test** | **Optimism** |
| --- | --- | --- | --- |
| **AUC ROC** | 0.9656 | 0.964 | 0.0016 |
| **Sommers D** | 0.9300 | 0.9268 | 0.0032 |
| **R2** | 0.7335 | 0.7271 | 0.0065 |
| **Intercept** | 0.0000 | -0.0147 | 0.0147 |
| **Slope** | 1.0000 | 0.9801 | 0.0199 |
| **Emax** | 0.0000 | 0.0069 | 0.0069 |

**Supplementary Table S9: Discrimination and classification accuracy (using an illustrative 50% model probability threshold) of Models 1-4, in comparison to clinical diagnosis and islet autoantibodies alone, by participant race/ethnicity.** *Using (for retained c-peptide) an illustrative ≥50% model probability cut off, provider diagnosis of Type 2 diabetes, or absence of islet-autoantibodies. Sensitivity and specificity are for retained c-peptide.

+ provider type with those a provider diagnosis other than Type 1 or 2 diabetes or unknown excluded

++ AUC ROC assessed using number of positive islet-autoantibodies, predictive value and accuracy assessed using any positive islet-autoantibody verses all negative.

Note that for islet autoantibodies AUC ROC, sensitivity, specificity and predictive values relates to inverted outcome (lower antibody number (AUC ROC) or negative islet-autoantibody status (other measures) indicating retained c-peptide).

| Ethnicity/Race | **Black** | | **Hispanic-White** | | **European-White** | |
| --- | --- | --- | --- | --- | --- | --- |
|  | **AUC ROC**  **(95% CI)** | **Overall accuracy* (95% CI)** | **AUC ROC**  **(95% CI)** | **Overall accuracy* (95% CI)** | **AUC ROC**  **(95% CI)** | **Overall accuracy* (95% CI)** |
| Routine measures  (Model 1) | 0.949 (0.930, 0.968)  n = 488 | 86.6  (83.3, 89.5) %  n = 488 | 0.937 (0.908, 0.964)  n = 422 | 88.4  (84.9, 91.2) %  n = 422 | 0.876  (0.835, 0.917)  n = 1937 | 95.8  (94.8, 96.7) %  n = 1937 |
| Routine measures + islet-autoantibodies (Model 2) | 0.971  (0.957, 0.985)  n = 488 | 93.1  (90.3, 95.1) %  n = 488 | 0.970 (0.947, 0.992)  n = 422 | 93.6  (90.8, 95.7) %  n = 422 | 0.937 (0.912, 0.961)  n = 1937 | 96.8  (95.9, 97.5) %  n = 1937 |
| Routine Measures + islet-autoantibodies and T1DGRS  (Model 3) | 0.969 (0.951, 0.986)  n = 350 | 93.1  (89.9, 95.6) %  n = 350 | 0.981 (0.957, 1.0)  n = 319 | 95.6 (92.7, 97.5) %  n = 319 | 0.962 (0.935, 0.989)  n = 1214 | 97.6  (96.6, 98.3) %  n = 1214 |
| Routine measures + T1DGRS  (Model 4) | 0.955 (0.934, 0.976)  n=350 | 89.4  (85.7, 92.4) %  n=350 | 0.964 (0.939, 0.989)  n=319 | 91.6  (87.9, 94.3) %  n=319 | 0.930 (0.893, 0.966)  n=1214 | 96.6  (95.4, 97.5) %  n=1214 |
| Healthcare provider diagnosis^+^ | 0.881  (0.852, 0.910)  n=481 | 88.6  (85.2, 91.3) %  n=481 | 0.906  (0.872, 0.940)  n=422 | 92.9  (90.0, 95.1) %  n=422 | 0.794 (0.747, 0.841)  n=1929) | 96.7  (95.8, 97.5) %  n=1929 |
| Islet-autoantibody number or status^++^ | 0.890 (0.863, 0.918)  n=488 | 87.9  (84.7, 90.7) %  n=488 | 0.929  (0.906 0.952)  n=422 | 90.5  (87.3, 93.1) %  n=422 | 0.843 (0.803, 0.884)  n=1937 | 87.4 (85.8, 88.8) %  n=1937 |

**Supplementary Figure S3: Model separation and calibration by race/ethnicity.** Model 1 (A - C), Model 2 (D - F), Model 3 (G - I) and Model 4 (J - L) by black, Hispanic-white, and European-white participants respectively. Left column: model probability by c-peptide outcome. Right Column: model calibration (development dataset): deciles of model predicted probabilities of retained c-peptide plotted against observed c-peptide outcome.

**A: Model 1 - Black participants (n=488) (45% retained c-peptide)**

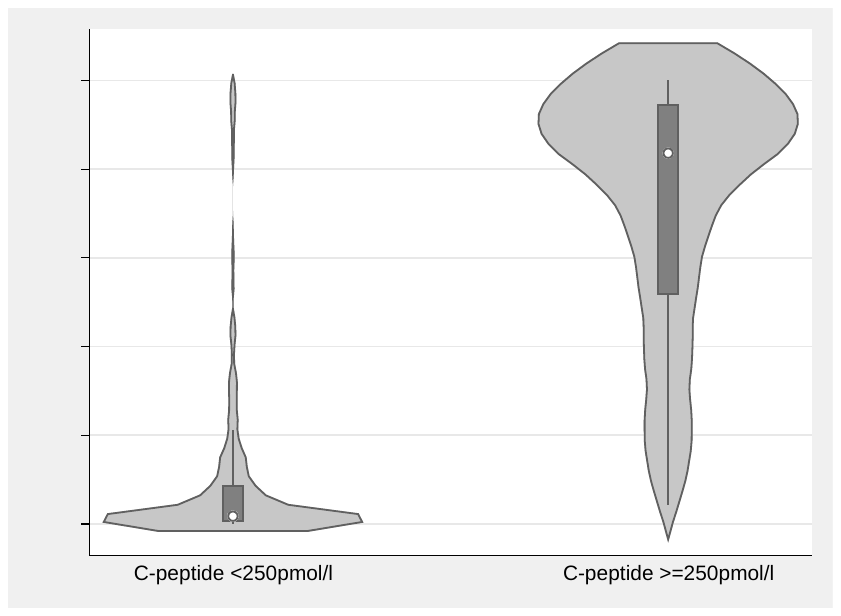

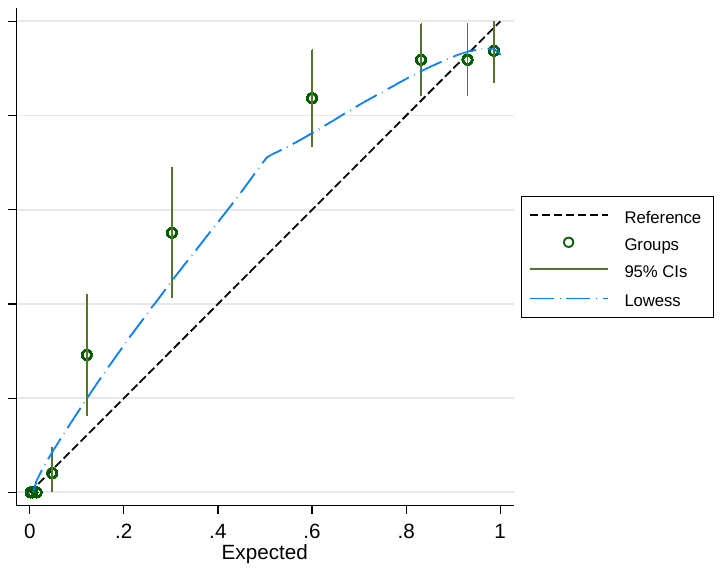

**B: Model 1 - Hispanic-white participants (n=422, 28% retained c-peptide)**

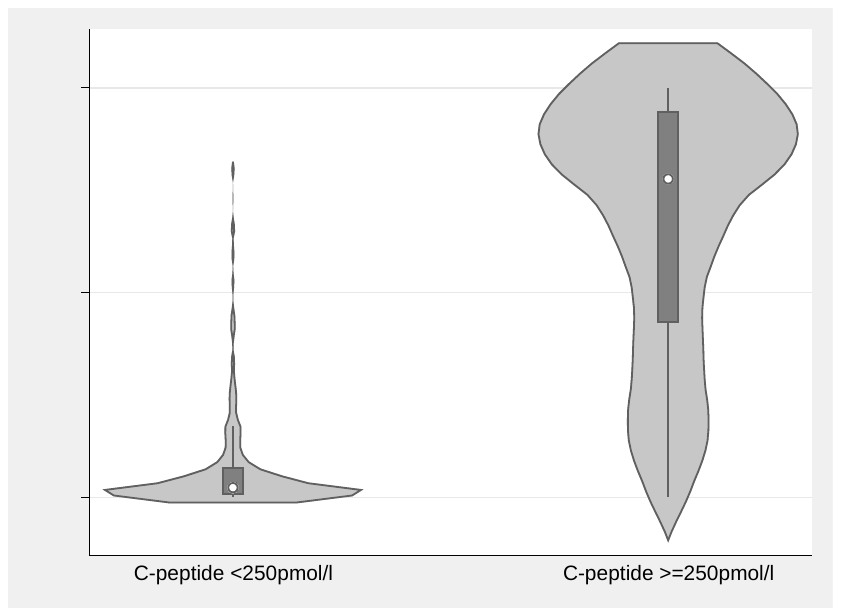

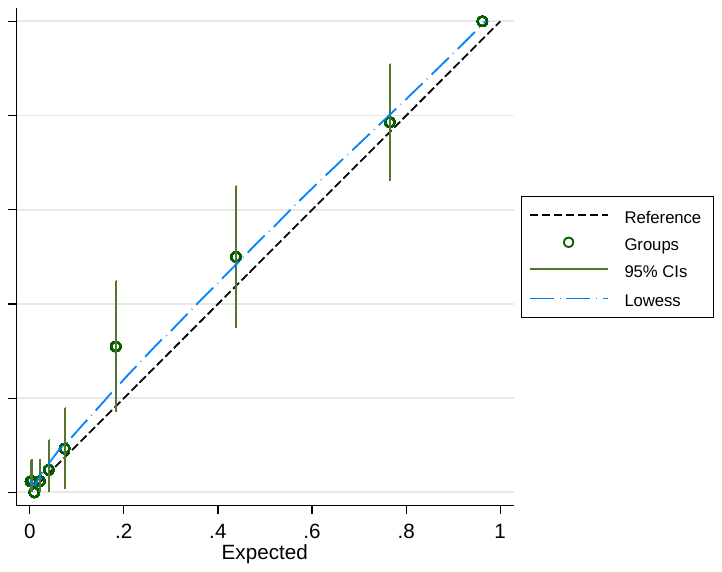

**C: Model 1 - European-white participants (n=1937, 6% retained c-peptide)**

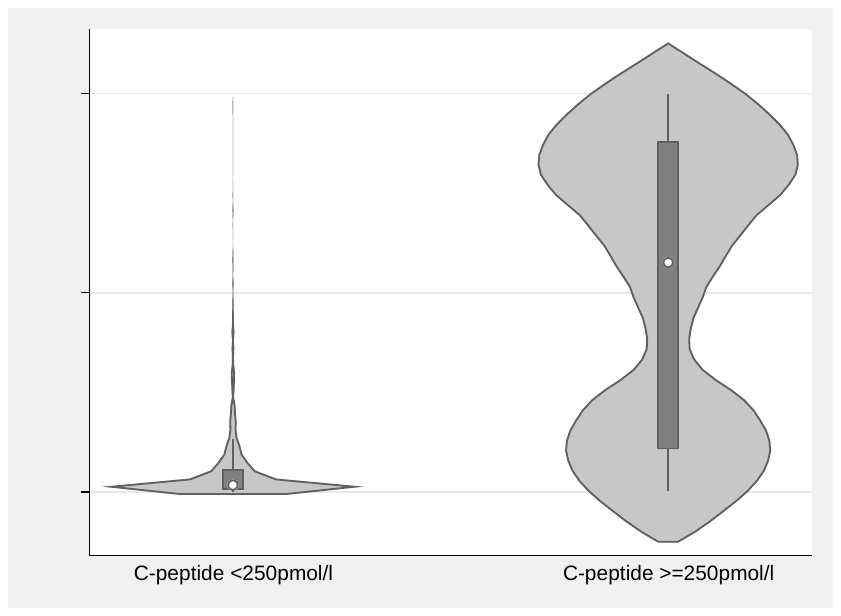

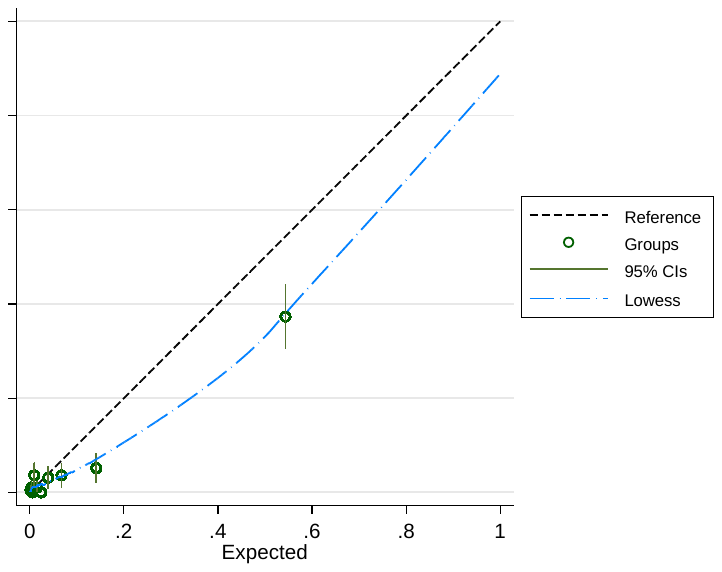

**D: Model 2 - Black participants (n=488, 45% retained c-peptide)**

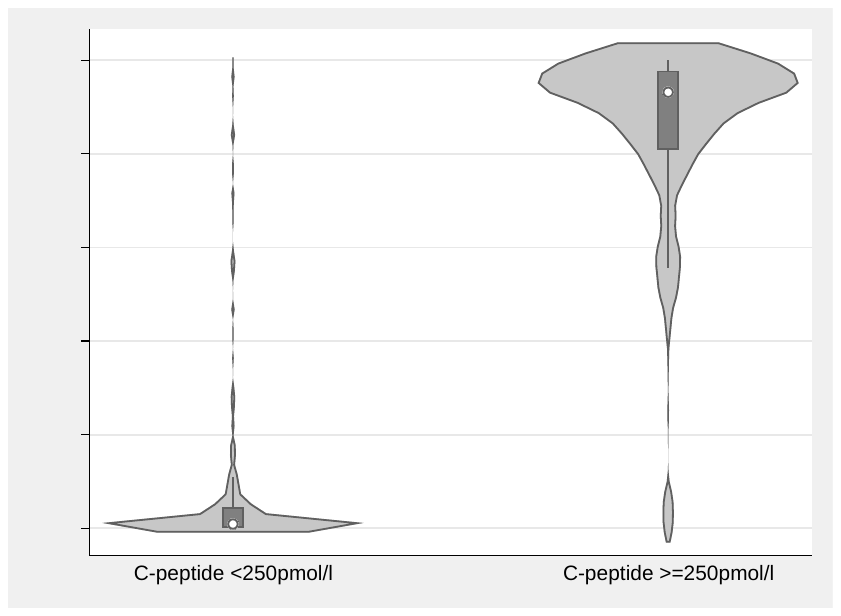

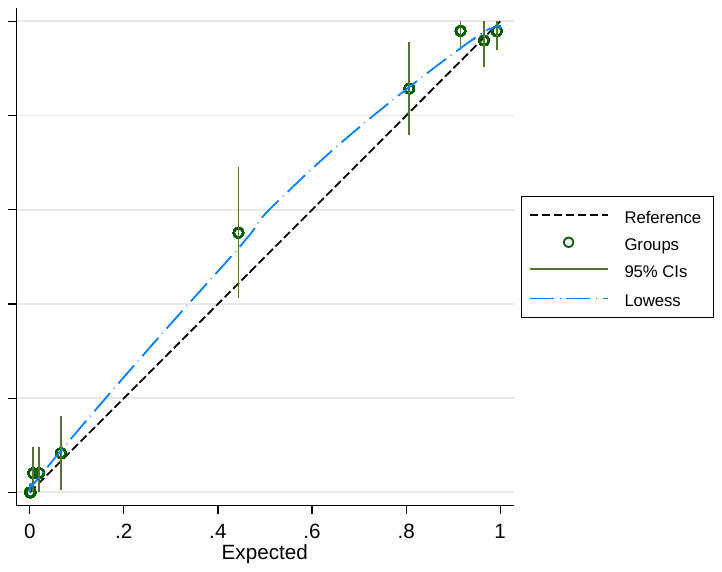

**E: Model 2 - Hispanic-White participants (n=422, 28% retained c-peptide)**

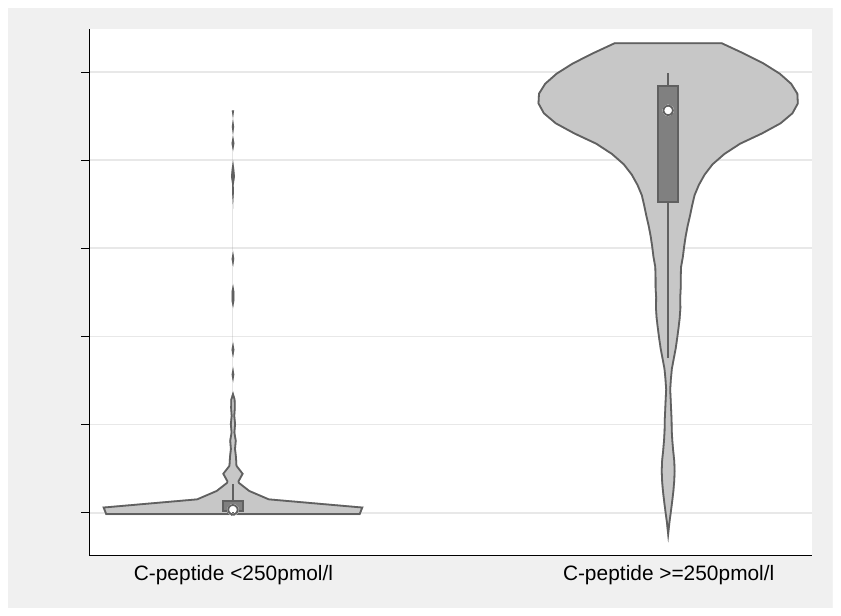

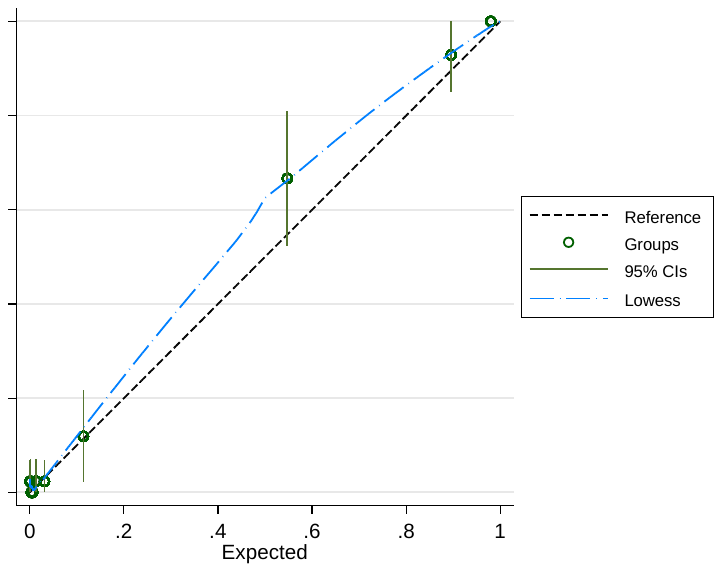

**F: Model 2 - European-White participants (n=1937, 6% retained c-peptide)**

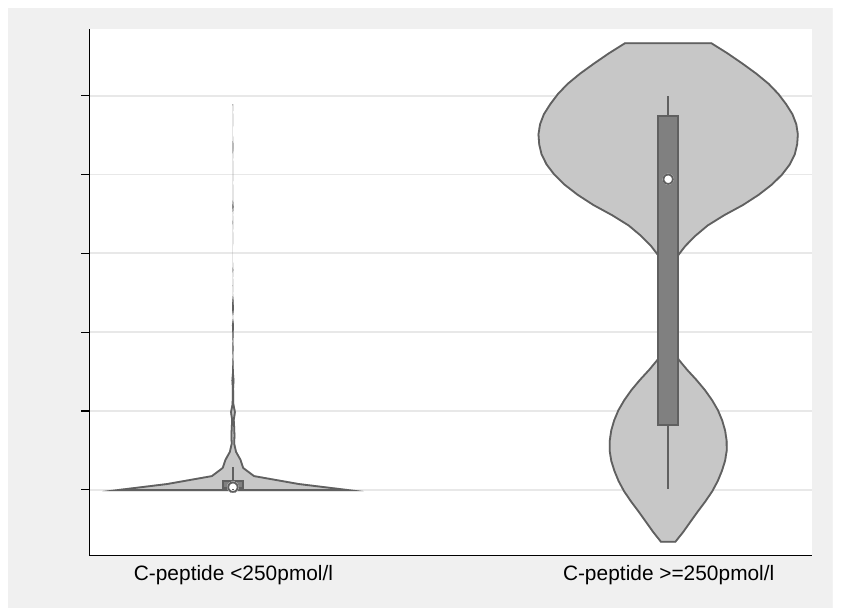

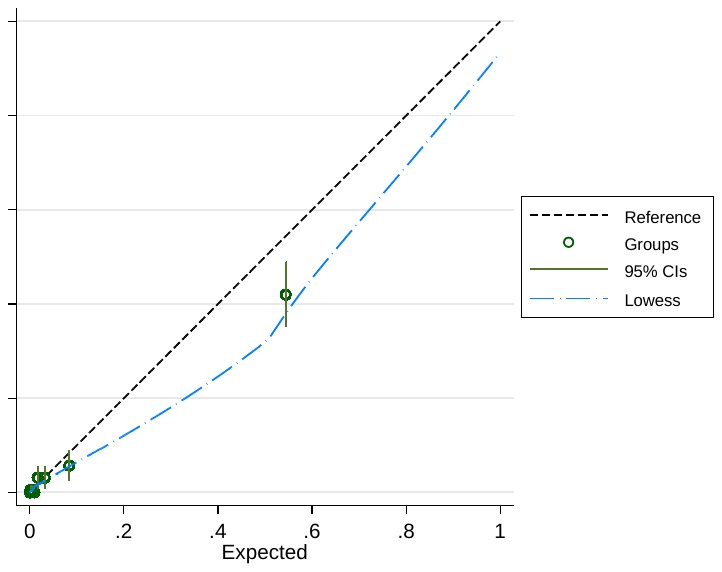

**G: Model 3 - Black participants (n=350, 39% retained c-peptide)**

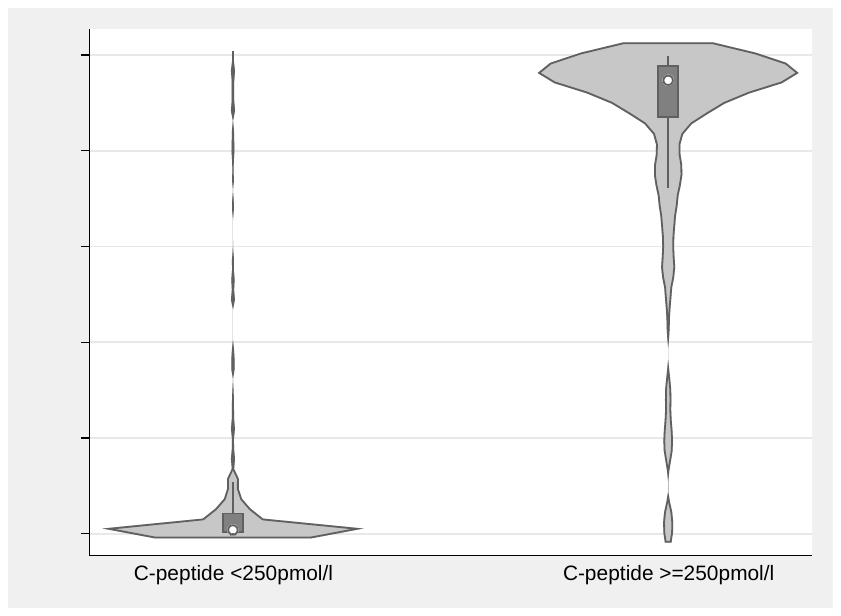

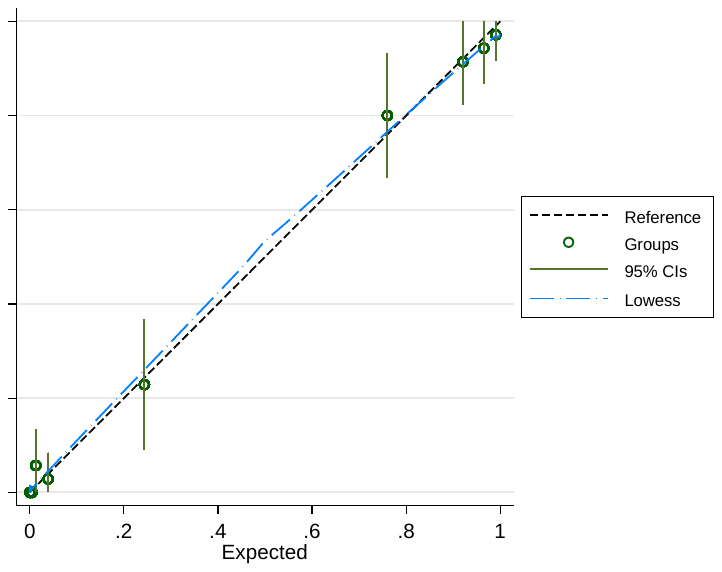

**H: Model 3 - Hispanic-white participants (n=319, 24% retained c-peptide)**

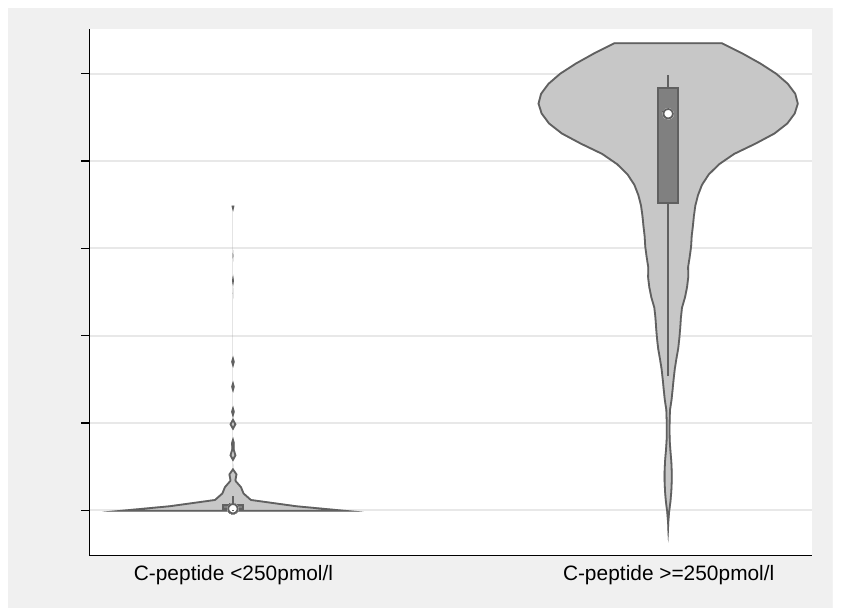

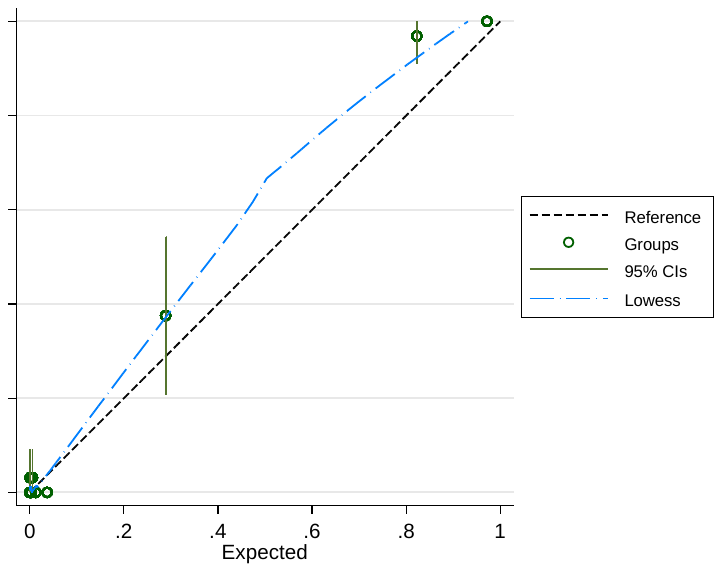

**I: Model 3 - European-White participants (n=1214, 6% retained c-peptide)**

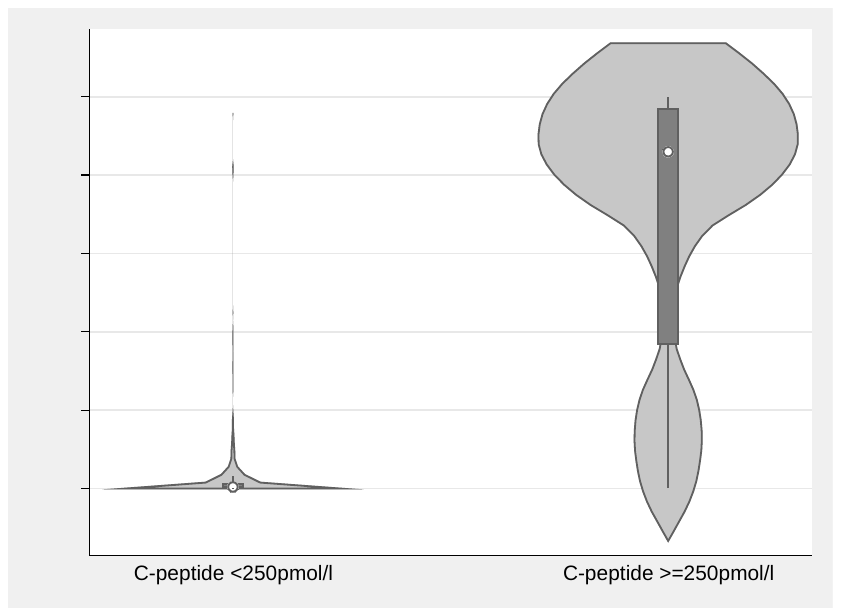

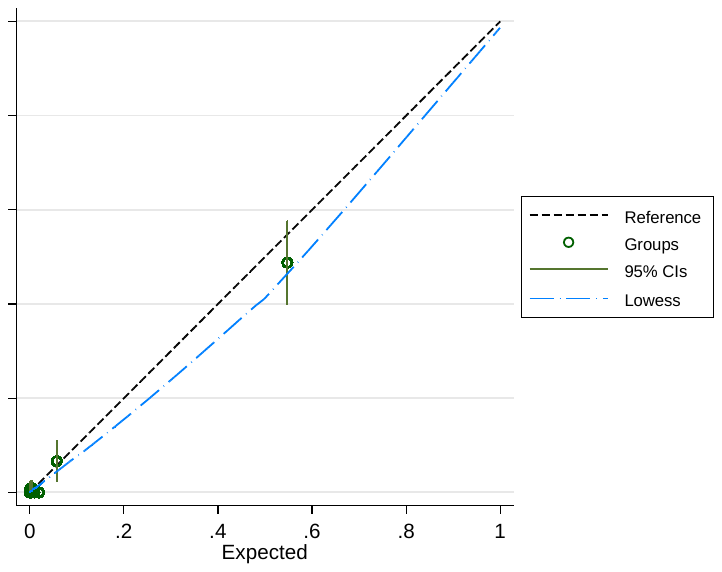

**J: Model 4 - Black participants (n=350, 39% retained c-peptide)**

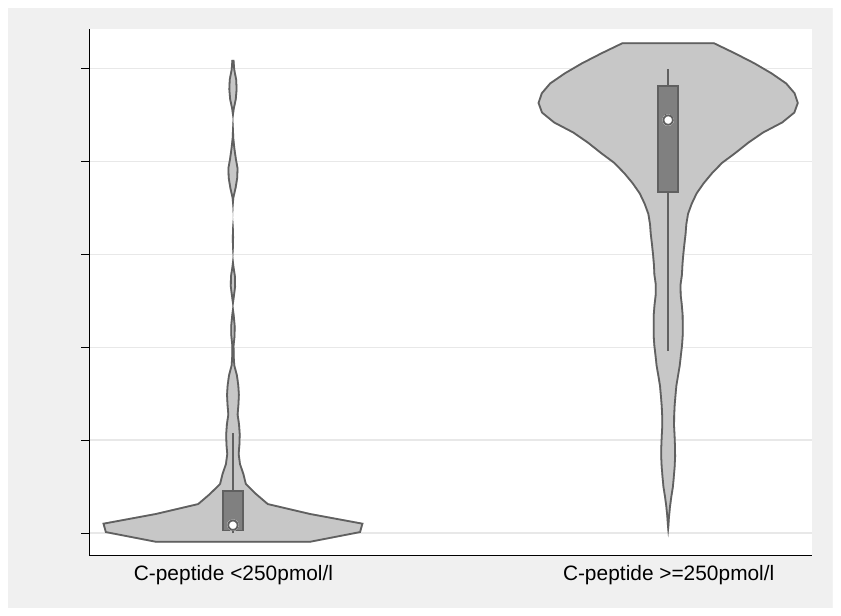

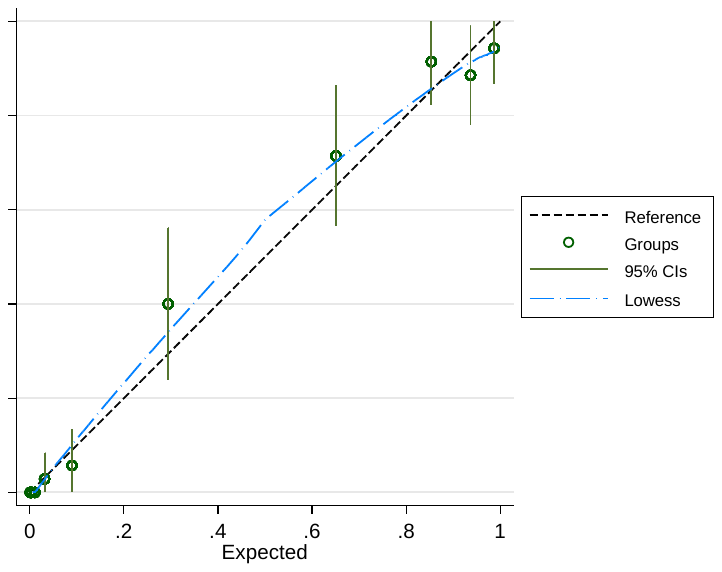

**K: Model 4 - Hispanic participants (n=319, 24% retained c-peptide)**

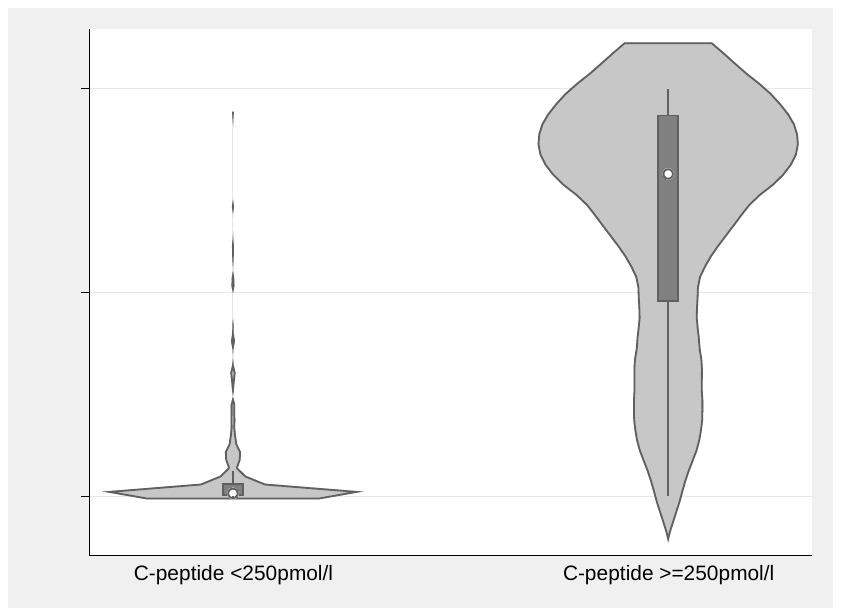

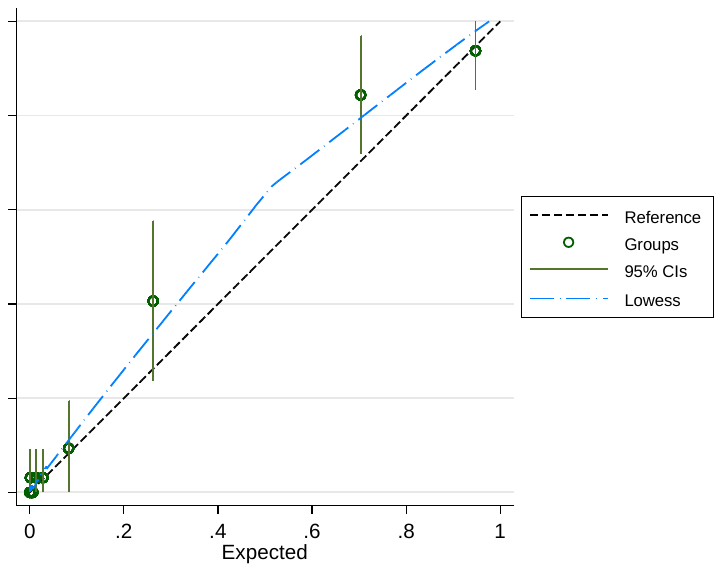

**L: Model 4 - European-White participants (n=1214, 6% retained c-peptide)**

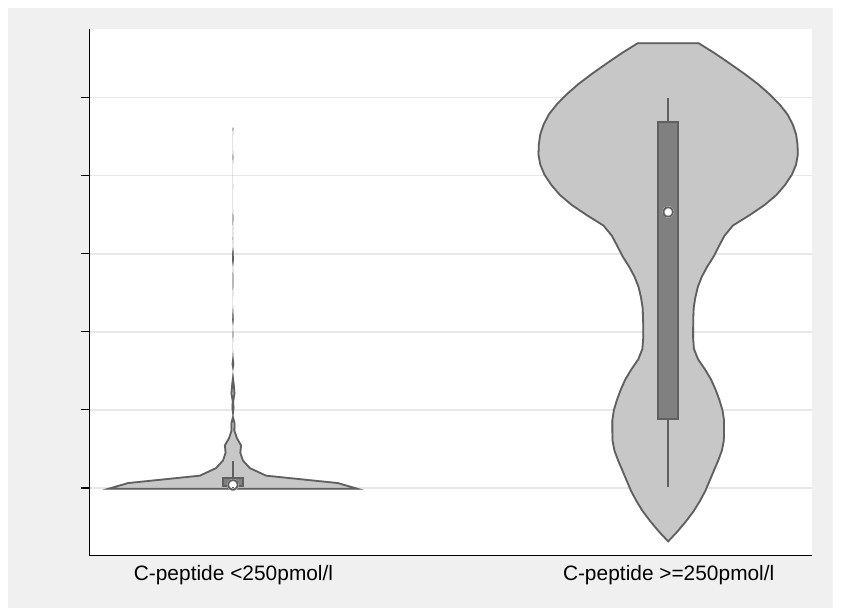

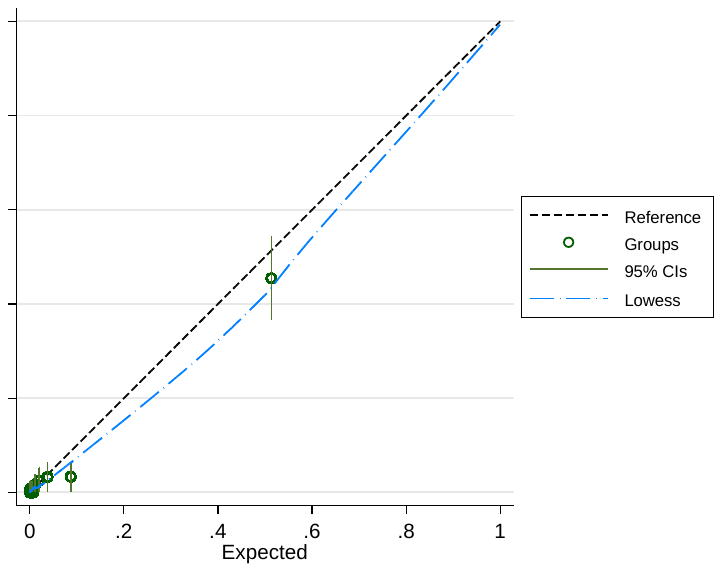

**Supplementary Table S10: Model performance with inclusion of race/ethnicity** (categorised as black, European-white, Hispanic-white, other based on prevalence in the SEARCH cohort). Other features are identical to Model 1-4 in the main manuscript.

**A:** **Discrimination and classification accuracy of Models 1-4 with the inclusion of race/ethnicity**

*Overall accuracy shows classification performance of an illustrative 50% Model cut off.

| **Features** | **AUC ROC**  **(95% CI)** | **Overall accuracy* (95% CI)** |
| --- | --- | --- |
| Routine measures + race/ethnicity  (Model 1R)  n = 2965 | 0.951 (0.938, 0.963) | 93.5 (92.5, 94.3) % |
| Routine measures + autoantibody status + race/ethnicity  (Model 2R)  n = 2965 | 0.973 (0.964, 0.981) | 95.8 (95.0, 96.5) % |
| Routine measures + autoantibody status + T1DGRS + race/ethnicity  (Model 3R)  n = 1918 | 0979 (0.969, 0.988) | 96.6 (95.7, 97.3) % |
| Routine measures + T1DGRS + race/ethnicity  (Model 4R)  n = 1918 | 0.965 (0.953, 0.977) | 94.7 (93.5, 95.6) % |

**B) Change in model performance with inclusion of race/ethnicity of Models 1-4 (in comparison with Models 1-4 without race/ethnicity)**

| **Model** | **n** | **Akaike's information criterion** | **Bayesian information criterion** | **Likelihood ratio test p value (comparison of Models with/without feature)** |
| --- | --- | --- | --- | --- |
| Model 1 without race/ethnicity | 2965 | 1196.836 | 1232.803 | <0.0001 |
| Model 1 with race/ethnicity | 2965 | 1107.547 | 1161.498 |  |
| Model 2 without race/ethnicity | 2965 | 836.7401 | 884.6971 | <0.0001 |
| Model 2 with race/ethnicity | 2965 | 808.8893 | 874.8303 |  |
| Model 3 without race/ethnicity | 1918 | 451.648 | 501.6794 | 0.03 |
| Model 3 with race/ethnicity | 1918 | 448.9676 | 515.676 |  |
| Model 4 without race/ethnicity | 1918 | 602.2007 | 641.1139 | 0.006 |
| Model 4 with race/ethnicity | 1918 | 595.7116 | 651.302 |  |

**Supplementary Figure S4: Separation and calibration plots for Models 1 - 4 (A - D) incorporating race/ethnicity.** Left column: model probability by c-peptide outcome. Right Column: model calibration (development dataset): deciles of model predicted probabilities of retained c-peptide plotted against observed c-peptide outcome.

**A: Model 1R - Routine measures only**

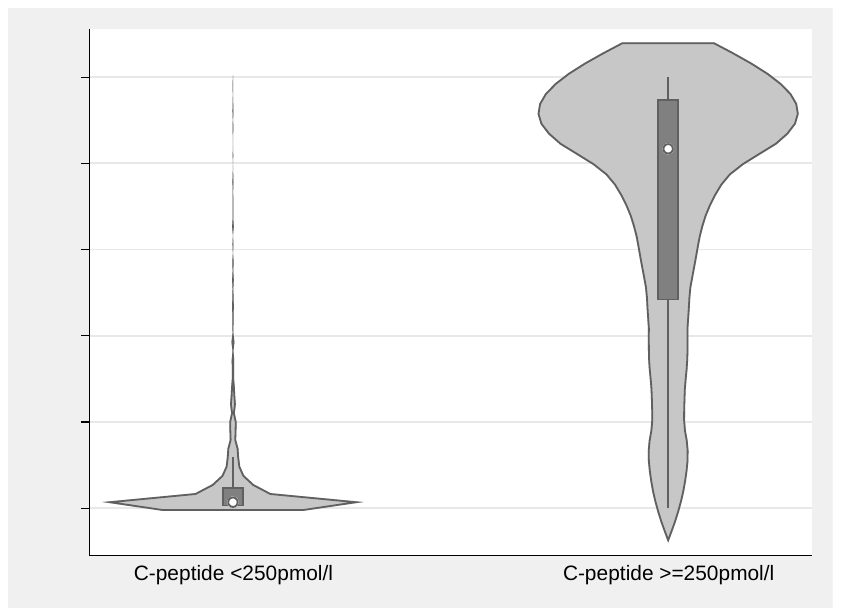

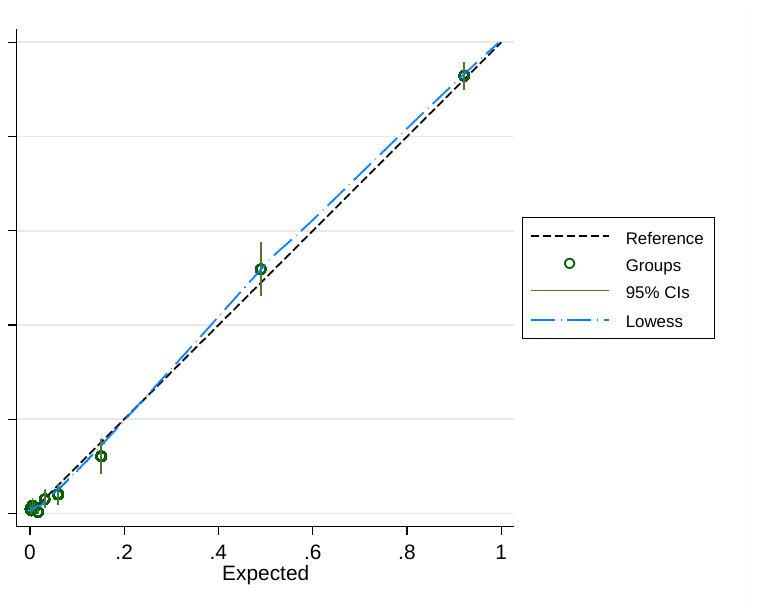

**B: Model 2R - Routine measures + islet-autoantibodies**

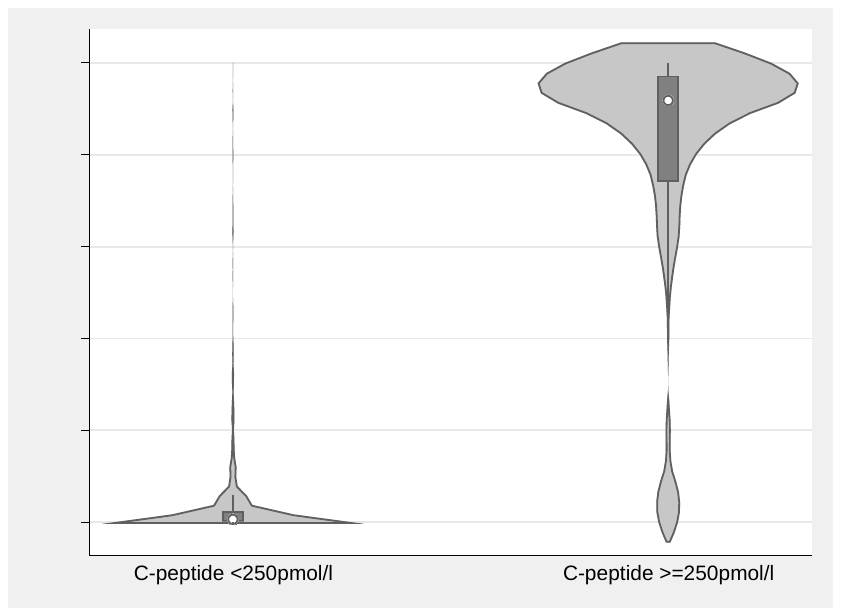

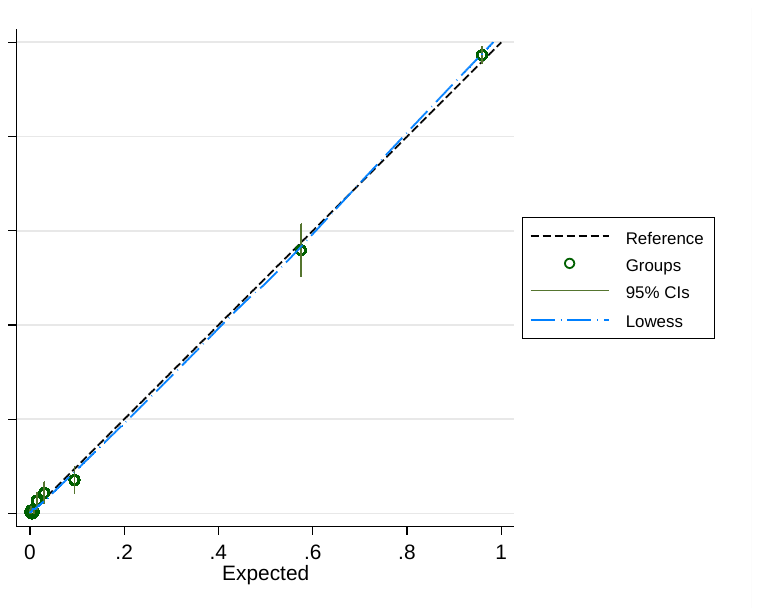

**C: Model 3R - Routine measures + Islet-autoantibodies + T1DGRS**

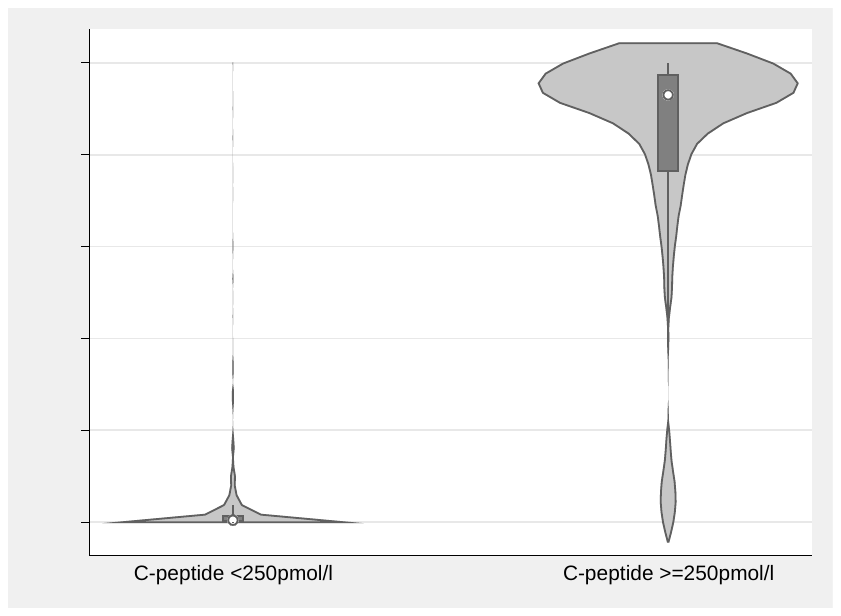

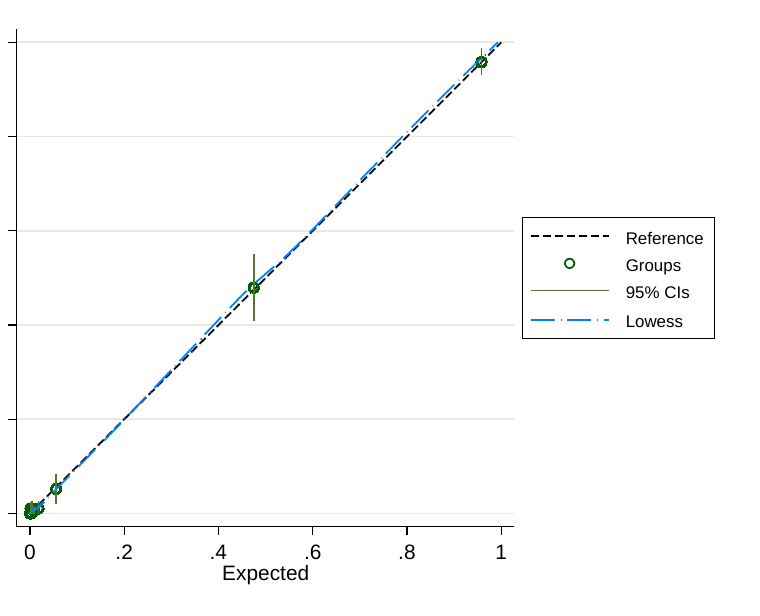

**D: Model 4R - Routine measures + T1DGRS**

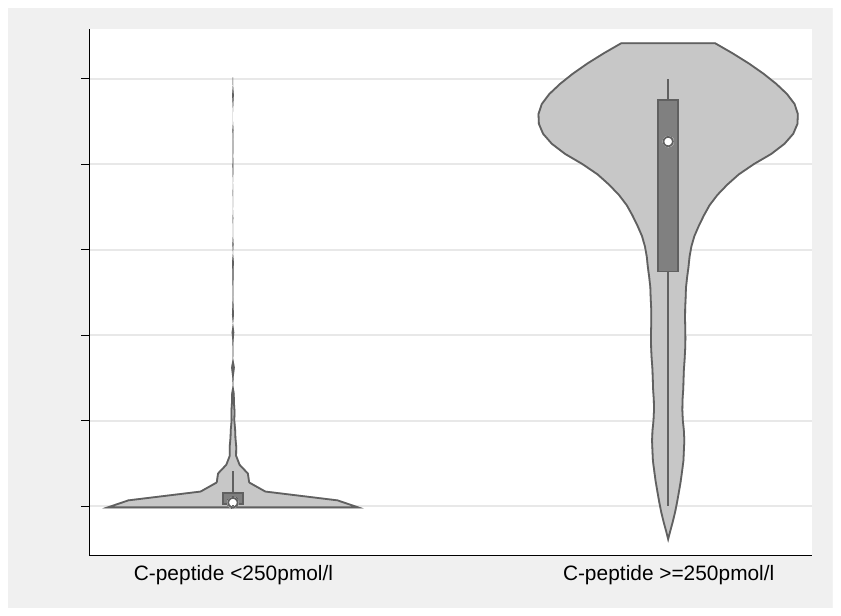

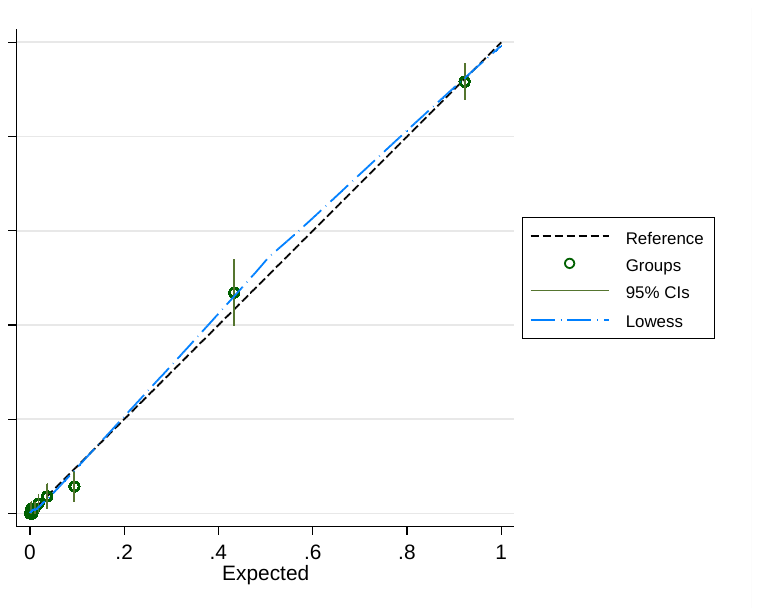

**Supplementary Table S10**: **Model performance in youth aged ≥ 10 years at diabetes diagnosis and obesity (BMI Z score >1.64)**.

Analysis for healthcare provider diagnosis and islet-autoantibody status limited to those with availability of Model 1 clinical features.

*Using (for retained c-peptide) a ≥50% Model probability cut off, provider diagnosis of Type 2 diabetes, or absence of islet-autoantibodies.

+ provider type (latest available) with those a provider diagnosis other than Type 1 or 2 diabetes excluded.

++ AUC ROC assessed using number of positive islet-autoantibodies, predictive value and accuracy assessed using any positive islet-autoantibody verses all negative.

Note that for islet autoantibodies AUC ROC, sensitivity, specificity, and predictive values relate to inverted outcome (lower antibody number (AUC ROC) or negative islet-autoantibody status (other measures) indicating retained C-peptide.

| **Features** | **AUC ROC**  **(95% CI)** | **Overall accuracy* (95% CI)** |
| --- | --- | --- |
| Routine measures only (Model 1) n=533 (c-peptide >250pmol/L n=377) | 0.855 (0.812, 0.892) | 82.4 (78.9, 85.5) % |
| Routine measures plus autoantibody status (Model 2) n = 533 (c-peptide >250pmol/L n=377) | 0.947(0.924, 0.969) | 91.2 (88.4, 93.4) % |
| Routine Measures plus negative Islet Autoantibodies and T1DGRS (Model 3)  n=333 (c-peptide >250pmol/L n=221) | 0.955 (0.929, 0.982) | 92.5 (89.1. 95.1) % |
| Routine measures + T1DGRS  (Model 4)  n=333 (c-peptide>250pmol/L n=221) | 0.912 (0.874, 0.949) | 86.3 (82.0 89.7) % |
| Healthcare provider diagnosis+  n=528 (c-peptide>250pmol/L n=339) | 0.849 (0.813, 0.884) | 87.1(84.0, 89.0) % |
| Islet-autoantibody number/status++  n=533 (c-peptide >250pmol/L n=377) | 0.887 (0.854 0.920) | 90.1 (88.0, 93.1) % |

**Supplementary Figure S6**: **Model separation and calibration in youth aged ≥ 10 years at diabetes diagnosis and obesity (BMI Z score >1.64) for Models 1 - 4 (A - D).** Left column: Model probability by c-peptide outcome. Right Column: Model calibration (development dataset) - deciles of model predicted probabilities of retained c-peptide plotted against observed c-peptide outcome.

**A: Model 1 -** **Routine measures only**

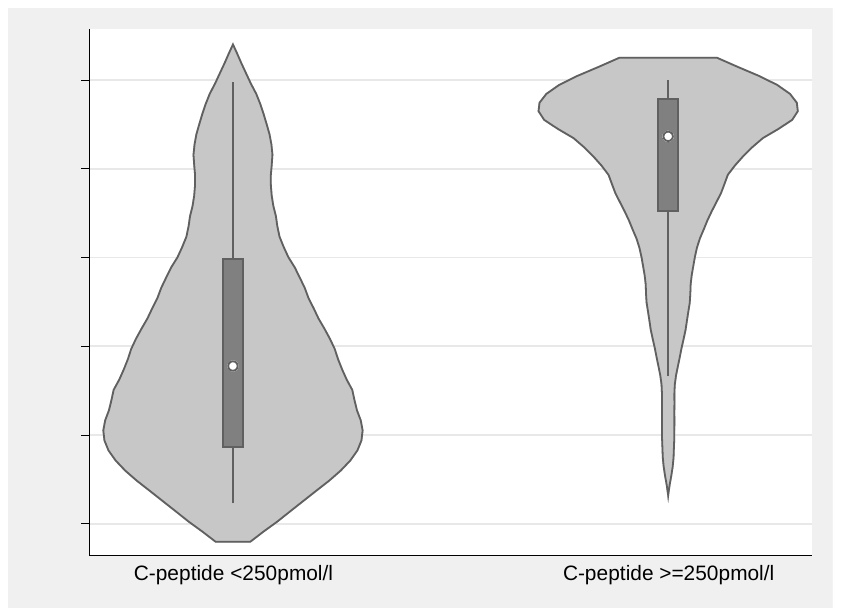

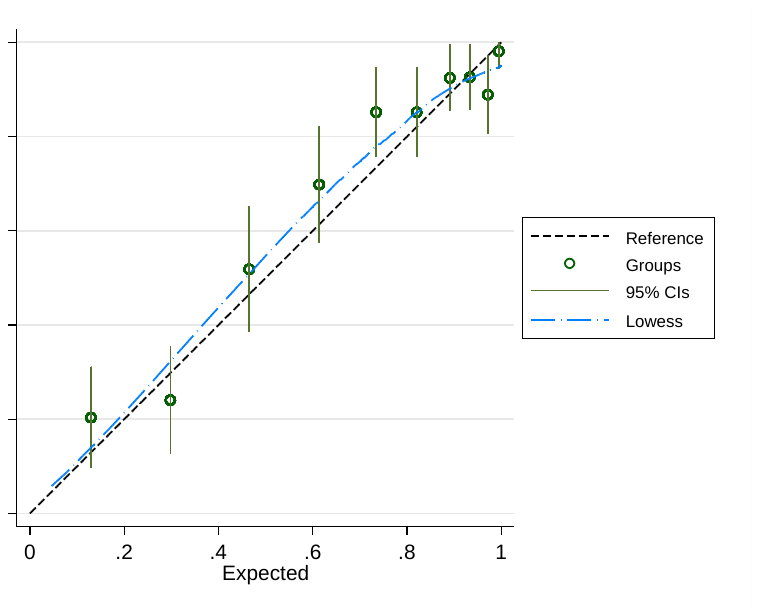

**B: Model 2 - Routine measures + islet-autoantibodies**

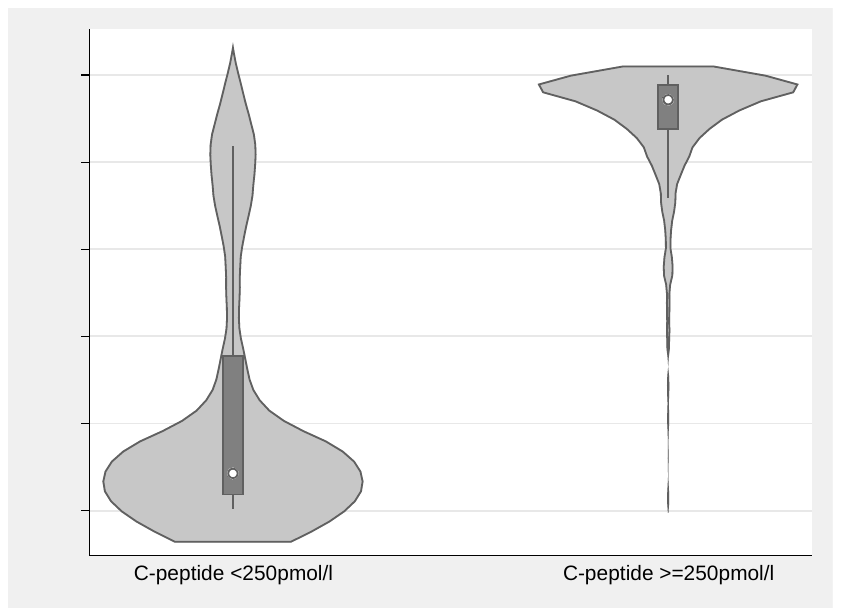

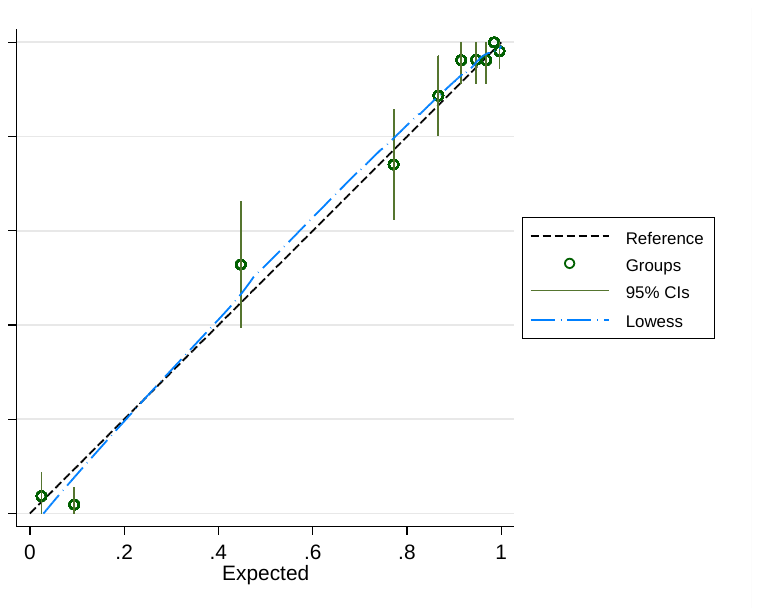

**C: Model 3** **- Routine measures + Islet-autoantibodies + T1DGRS**

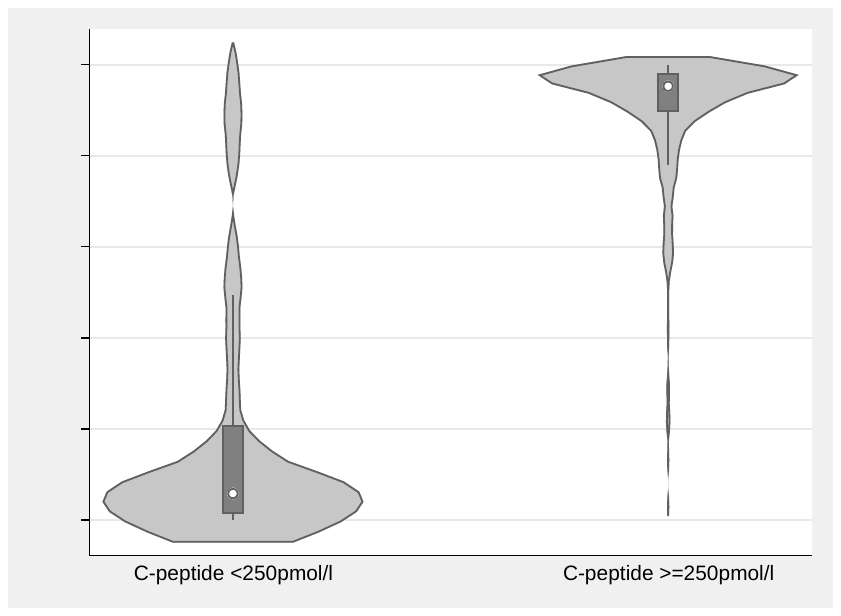

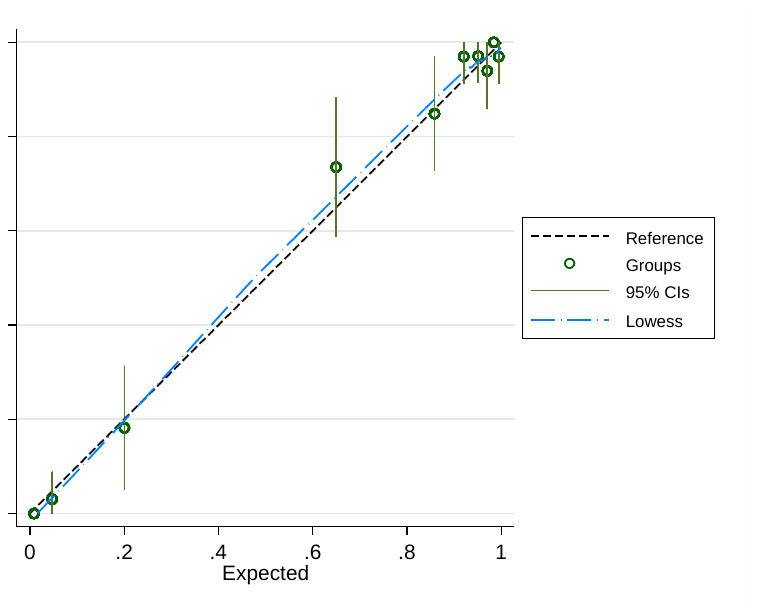

**D: Model 4 - Routine measures + T1DGRS**

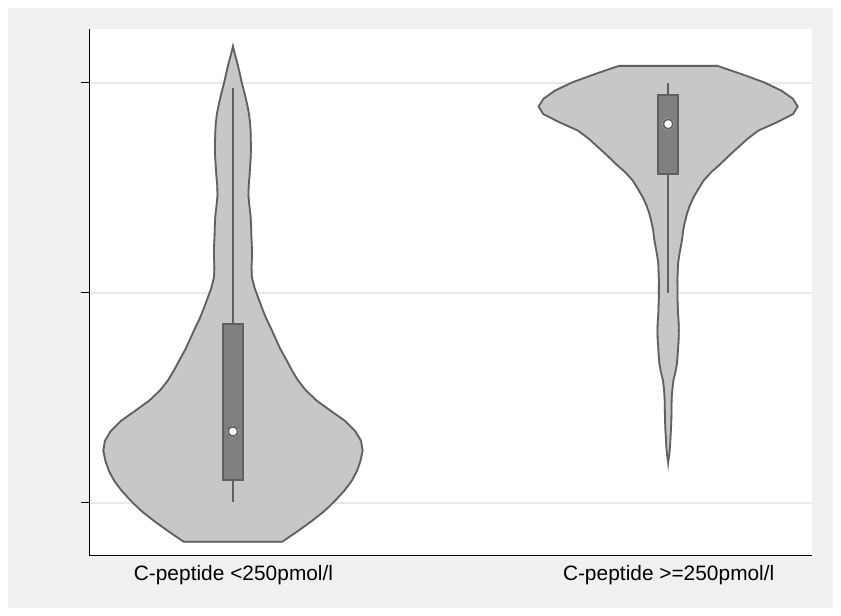

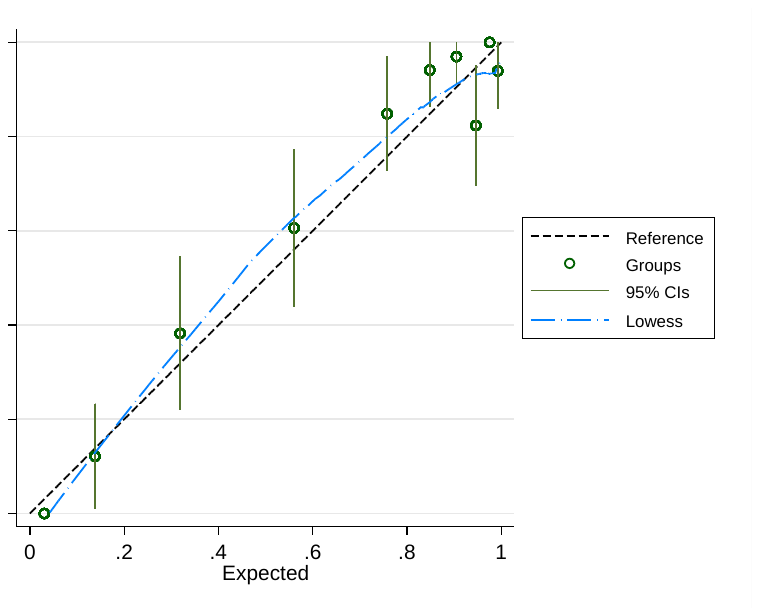

**Supplementary Figure S7: Model 2 and 3 separation and calibration in islet-autoantibody negative (A - B) and positive (C - D) participants.**

Left column: Model probability by c-peptide outcome. Right Column: Model calibration (development dataset) - deciles of model predicted probabilities of retained c-peptide plotted against observed c-peptide outcome.

**A: Model 2 - Islet-autoantibody negative participants**

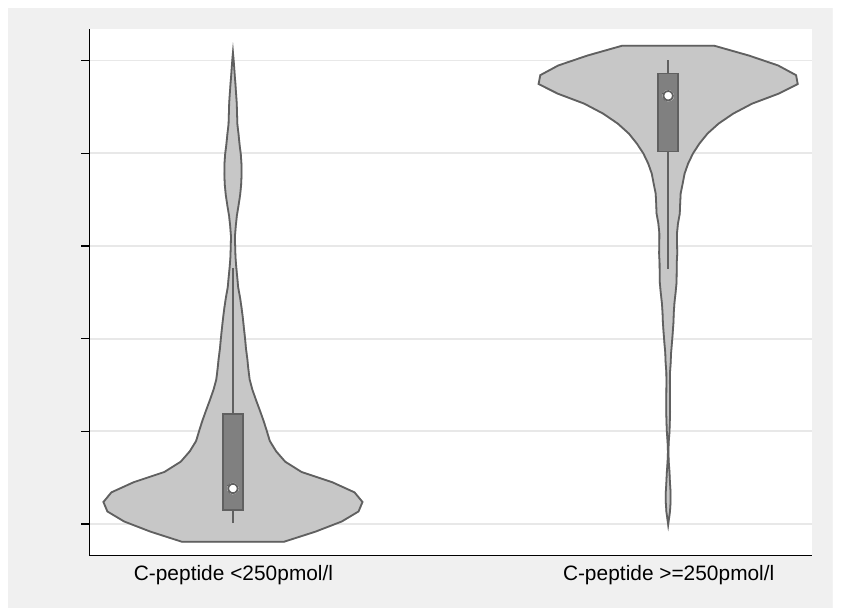

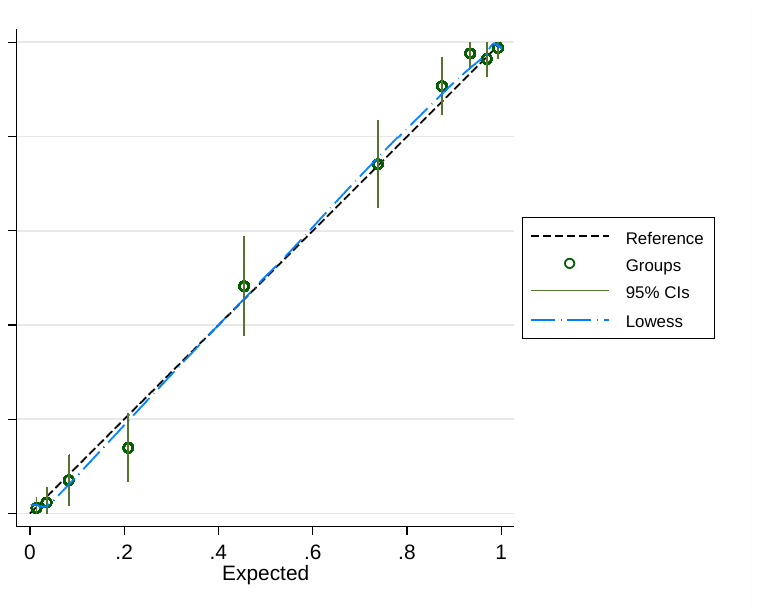

**B: Model 3 - Islet-autoantibody negative participants**

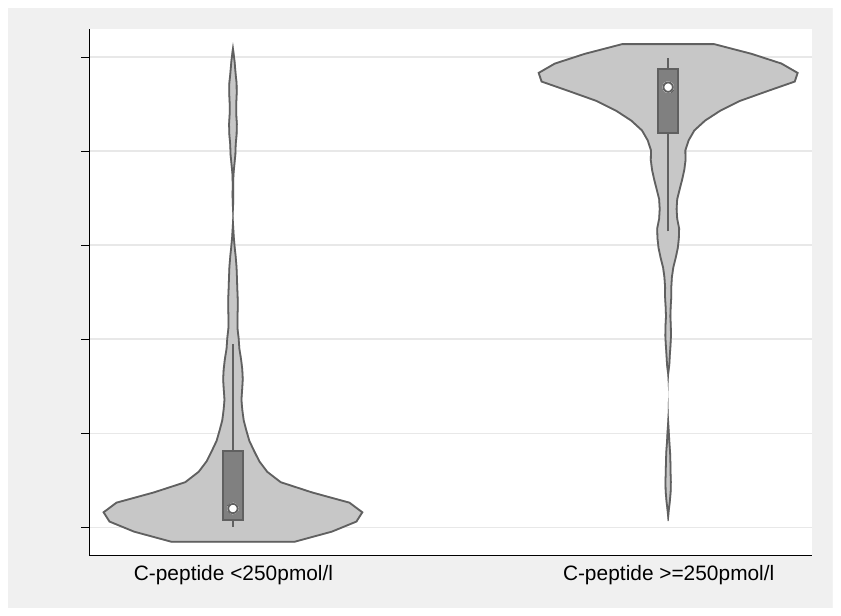

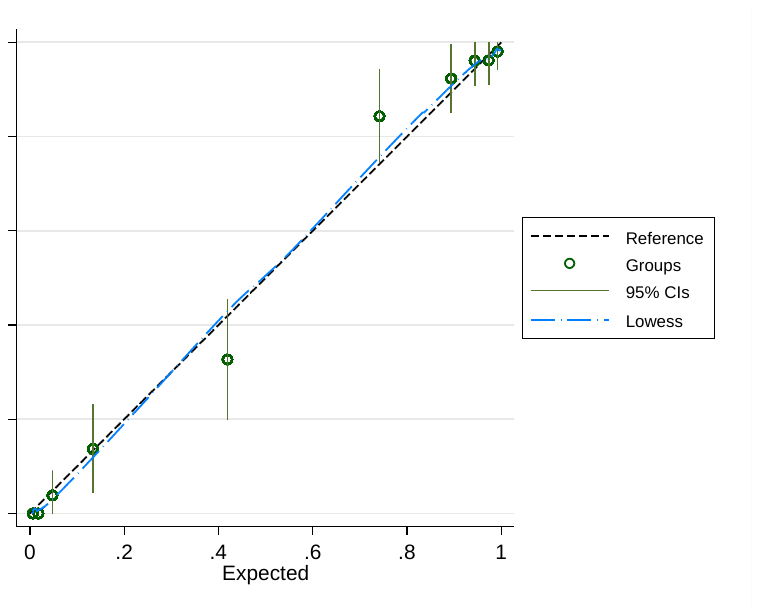

**C: Model 2 - Islet autoantibody positive participants**

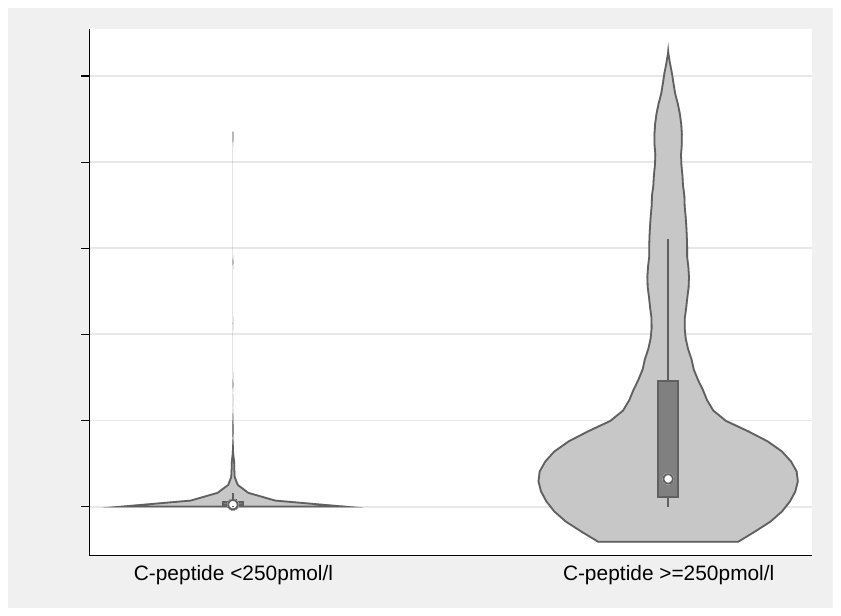

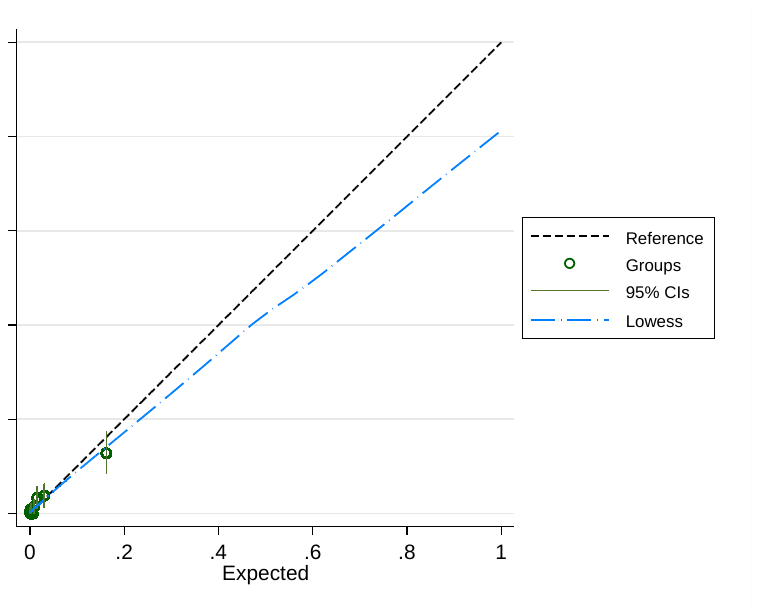

**D: Model 3 - Islet Autoantibody positive participants**

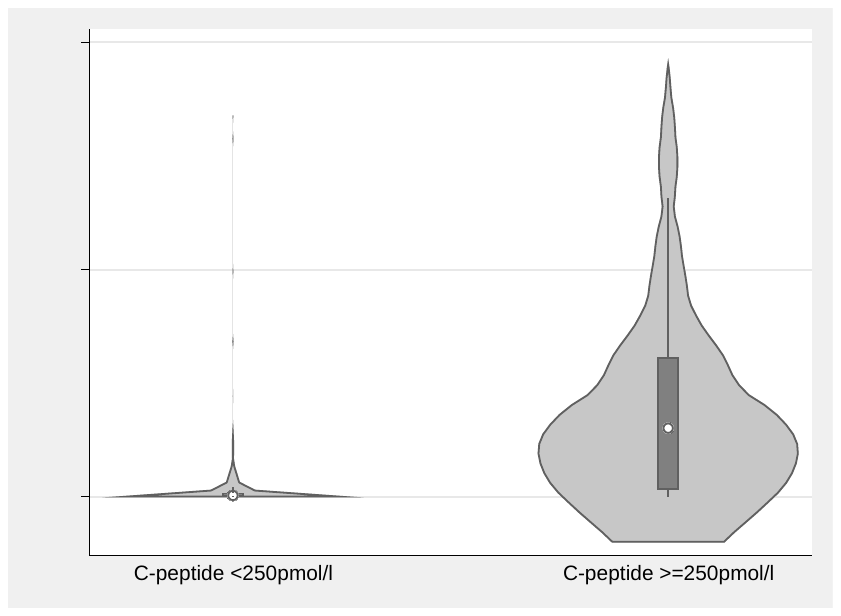

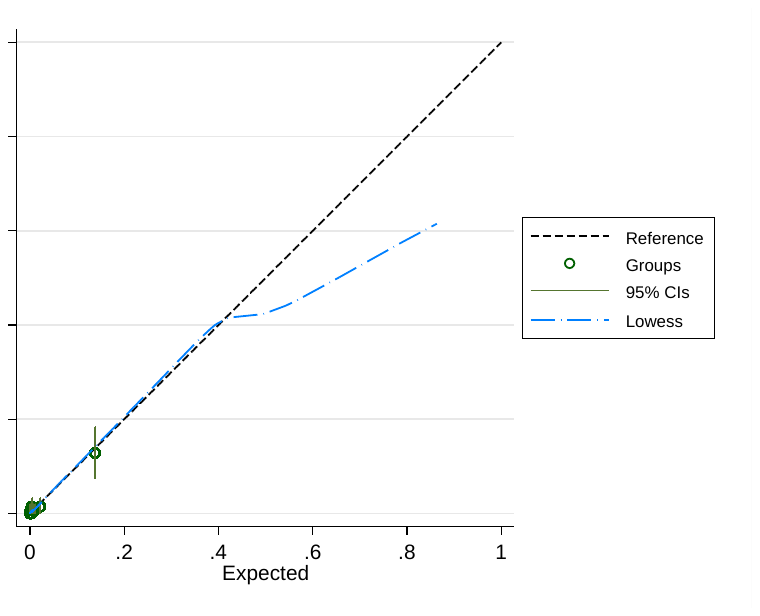

**Supplementary Table S11:** **Discrimination (AUC ROC) of models for retained c-peptide within subtype of diabetes defined by the SEARCH study aetiology-based classification system.**

AA = Islet autoantibody. IR = Insulin resistance.

Total number (total with retained c-peptide) for models without T1DGRS (Models 1-2) and with T1DGRS (Models 3-4): AA+ve IR-ve: 1350(17) and 892(9), AA+ve IR+ve: 442(26) and 297 (12), AA-ve IR-ve: 139(32) and 80(19), AA-ve IR+ve: 401(348) and 228(196).

| **Model** | **AA+ve IR-ve** | **AA+ve IR+ve** | **AA-ve IR-ve** | **AA-ve IR +ve** |
| --- | --- | --- | --- | --- |
| Model 1 | 0.850 (0.760, 0.941) | 0.853 (0.772, 0.934) | 0.861 (0.792, 0.941) | 0.899 (0.851, 0.948) |
| Model 2 | 0.850 (0.759, 0.941) | 0.869 (0.784, 0.954) | 0.865 (0.789, 0.939) | 0.896 (0.847, 0.944) |
| Model 3 | 0.822 (0.680, 0.964) | 0.933 (0.876, 0.990) | 0.909 (0.838, 0.979) | 0.866 (0.786, 0.946) |
| Model 4 | 0.819 (0.684, 0.955) | 0.901 (0.832, 0.984) | 0.899 (0.850, 0.947) | 0.863 (0.781, 0.945) |

**Supplementary Table S12: The impact of adding Znt8 islet-autoantibody testing on Model 2 and 3 (Models with GAD and IA2 islet-autoantibodies), limited to participants with all three islet-autoantibodies available (n = 2767 (Model 2) and 1888 (Model 3)).**

**A: Impact of adding Znt8 islet-autoantibody to discrimination performance (AUC ROC) of Models 2 and 3**

| **Model** | **AUC ROC With Znt8 (95% CI)** | **AUC ROC without Znt8 (95% CI)** |
| --- | --- | --- |
| Model 2 | 0.975 (0.967, 0.983) | 0.973 (0.965, 0.981) |
| Model 3 | 0.980 (0.970, 0.989) | 0.978 (0.967, 0.988) |

**B: Impact of adding Znt8 islet-autoantibody to model performance (Likelihood ratio test, Akaike's information criterion (AIC) and Bayesian information criterion (BIC))** **of Models 2 and 3**

Note a lower number on AIC and BIC reflects better model fit

| **Model** | **n** | **Akaike's information criterion** | **Bayesian information criterion** | **Likelihood ratio test p value (comparison of Models with/without feature)** |
| --- | --- | --- | --- | --- |
| Model 2 without Znt8 | 2767 | 783.0643 | 830.4685 | <0.001 |
| Model 2 with Znt8 | 2767 | 747.8425 | 801.1721 |  |
| Model 3 without Znt8 | 1888 | 446.688 | 496.5774 | <0.001 |
| Model 3 with Znt8 | 1888 | 430.0972 | 485.5299 |  |

**Supplementary Tables S13: The impact of adding a type 2 diabetes genetic risk score (T2DGRS) on Model 3 and 4 (Models with T1DGRS), limited to participants with T2DGRS available (n = 1918).**

**A:** **Impact of adding T2DGRS to discrimination performance (AUC ROC) of Models 3 and 4**

| **Model** | **AUC ROC With T2DGRS (95% CI)** | **AUC ROC without T2DGRS (95% CI)** |
| --- | --- | --- |
| Model 3 | 0.979 (0.969, 0.989) | 0.978 (0.968, 0.988) |
| Model 4 | 0.965 (0.953, 0.977) | 0.966 (0.953, 0.977) |

**B:** **Impact of adding T2DGRS to model performance (Likelihood ratio test, Akaike's information criterion (AIC) and Bayesian information criterion (BIC))** **of Models 3 and 4**

Note a lower number on AIC and BIC reflects better model fit

| **Model** | **n** | **Akaike's information criterion** | **Bayesian information criterion** | **Likelihood ratio test p value (comparison of Models with/without feature)** |
| --- | --- | --- | --- | --- |
| Model 3 without T2DGRS | 1918 | 451.648 | 501.6794 | 0.03 |
| Model 3 with T2DGRS | 1918 | 448.9809 | 504.5712 |  |
| Model 4 without T2DGRS | 1918 | 602.2007 | 641.1139 | <0.001 |
| Model 4 with T2DGRS | 1918 | 587.575 | 632.0473 |  |

**Supplementary Table S14: Performance (AUC ROC) of Models without HDL and waist circumference (Model 1 and 2 n = 2966, Model 3 and 4 n = 1918)**

| **Model** | **AUC ROC (95% CI)** |
| --- | --- |
| Model 1 | 0.939 (0.826, 0.951) |
| Model 2 | 0.970 (0.961, 0.978) |
| Model 3 | 0.976 (0.966, 0.987) |
| Model 4 | 0.960 (0.947, 0.973) |

**Supplementary Table S15: Model performance (AUC ROC) using a 30 SNP type 1 diabetes genetic risk score (T1DGRS-1), in place of the 67 SNP score (T1DGRS-2) reported in the main analysis, limited to participants with both scores available. N = 1905 for all.**

|  | **T1DGRS2**  **AUC ROC (95% CI)** | **T1DGRS1**  **AUC ROC (95% CI)** |
| --- | --- | --- |
| Model 3 with T1DGRS -1 | 0.979 (0.970, 0.989) | 0.979 (0.970, 0.989) |
| Model 4 with T1DGRS-1 | 0.966 (0.954, 0.977) | 0.962 (0.951, 0.974). |

**Supplementary Table S16: Model coefficients.**

To convert output (log odds) to probability use the formula exp(log odds)/(1+exp(log odds)).

Sex coded as 0 (female) 1 (Male). Units for continuous variables are years for Age (of diagnosis), Kg/m^2^ for BMI, cm for waist circumference, mg/dl for HDL,. T1DGRS1 and T1DGRS2 are specific to the scores described in the methods section.

| **Model** | **Equation** |
| --- | --- |
| Main Models | |
| Model 1 (Clinical features only) | -6.843912 + (-0.8700783*sex) + (0.0739423*age) + (0.1592899 *BMI) + (-0.0529831* HDL) + (0.0368719 *waist circumference) |
| Model 2 (Clinical features + GAD + IA2) | -4.502606 + (-0.9339223 *sex) + (0.1345481*age) + (0.138903 *BMI) + (-0.0435481* HDL) + (0.0209935 *waist circumference) + (-2.856424 if one positive antibody, -3.963131 if two positive antibodies) |
| Model 3 (Clinical features + GAD + IA2 + T1DGRS2) | - 0.1216426 + (-0.7741532*sex) + (0.1542869*age) + (0.108483 *BMI) + (-0.0369311* HDL) + (0.0177613 *waist circumference) + (-2.725245 if one positive antibody, -3.751959 if two positive antibodies) + (-0.3859143* T1DGRS2) |
| Model4 (Clinical features + T1DGRS2) | 0.7560816 + (-0.8380161*sex) + (0.0896181*age) + (0.1171035*BMI) + (-0.0481782 * HDL) + (0.0292977*waist circumference) + (-0.5689743 * T1DGRS2) |
| Additional Model variants | |
| Model 1 with Race/ethnicity | -5.100375 + (-0.6953544 *sex) + (0.0751763 *age) + (.1060905*BMI) + (-0.0569685* HDL) + (0.0443439 *waist circumference) + (0 if Black, -0.4914if Hispanic White, -1.712099 if Non-Hispanic White) |
| Model 2 with Race/ethnicity | -3.293692 + (-0.7541725*sex) + (0.1260244*age) + (0.1060389 *BMI) + (-0.0453029 * HDL) + (0.0243402*waist circumference) + (-2.71073 if one positive antibody, -3.747755 if two positive antibodies) + (0 if black, -0.1427544 if Hispanic White, -1.185805 if Non-Hispanic White) |
| Model 3 with Race/ethnicity | -0.0591628 + (-0.6925918*sex) + (0.1476899 *age) + (0.0979397 *BMI) + (-0.0358216 * HDL) + (0.0201225 *waist circumference) + (--2.769734 if one positive antibody, -3.752584 if two positive antibodies) + (-0.355778 * T1DGRS2) + (0 if black, 0.5898827 if Hispanic White, -0.432062 if Non-Hispanic White) |
| Model 4 with Race/ethnicity | -0.8927357 + (-0.7583089 *sex) + (0.0824723 *age) + (0.1025489 *BMI) + (0.0490835* HDL) + (0.0319338 *waist circumference) + (-0.5410167 * T1DGRS2) + (0 if black, 0.3706463 if Hispanic White, 0.5754588 if Non-Hispanic White) |
| Model 2 with GAD, IA2 and Znt8 measured islet autoantibodies | (-4.297873) + (-0.8318433*sex) + (0.1532406*age) + (0.1328409*BMI) + (-0.0426679*HDL) + (0.0196024*waist circumference) + (-2.882518 if one positive antibody, -3.571513 if two positive antibodies, -4.528076 if three positive antibodies) |
| Model 3 with GAD, IA2 and Znt8 measured islet autoantibodies | 0.2338514 + (-0.6617823*sex) + (0.163667*age) + (0.0986245*BMI) + (-0.037171*HDL) + (0.0164291*waist circumference) + (-2.79436 if one positive antibody, -3.402645 if two positive antibodies, -4.468159 if three positive antibodies) + (-0.3507495*T1DGRS2) |
| Model 3 with T1DGRS1 | 2.395272 + (-0.8169852 *sex) + (0.1625459*age) + (0.1310264 *BMI) + (--0.0369966*HDL) + (0.0085764 *waist circumference) + (-2.738345 if one positive antibody, -3.945259 if two positive antibodies) + (-25.12357 *T1DGRS1) |
| Model 4 with T1DGRS1 | 3.039244 + (-.07593889 *sex) + (0.0835448 *age) + (0.142629 *BMI) + (-0.0454263 *HDL) + (0.0240089 *waist circumference) + + (-34.80119 *T1DGRS1) |
| Model 1 without waist circumference and HDL | -9.660739 + (-0.6602995*sex) + (0.1218207*age) + (0.2688966*BMI) |
| Model 2 without waist circumference and HDL | -7.065322 + (-0.740672*sex) + (0.1723298*age) + (0.2084922*BMI) + (-2.93537 if one positive antibody, 4.169799 if two positive antibodies) |
| Model 3 without waist circumference and HDL | -1.947001 + (-0.6368531*sex) + (0.1911258*age) + (0.1627751*BMI) + (-2.809559 if one positive antibody, 3.960101 if two positive antibodies) + (-0.390164*T1DGRS2) |
| Model 4 without waist circumference and HDL | -1.723943 + (-0.6310751*sex) + (0.1329611*age) + (0.2035865*BMI) + (-0.5830442*T1DGRS2) |
| Model 2 with ethnicity, GAD, IA2 and Znt8 measured islet autoantibodies | -3.279001 + (-0.6915499 *sex) + (0.1403291 *age) + (0.1073039 *BMI) + (-0.0444286 * HDL) + (0.0220058 *waist circumference) + (--2.786631 if one positive antibody, -3.337842 if two positive antibodies, -4.307275 if three positive antibodies) + (0 if black, -0.028466 if Hispanic White, -0.9822102 if Non-Hispanic White) |
| Model 3 with ethnicity, GAD, IA2 and Znt8 measured islet autoantibodies | 0.0072505 + (-0.6085475*sex) + (0.1547811 *age) + (0.093661*BMI) + (-0.0349692* HDL) + (0.0178491 *waist circumference) + (-2.821309 if one positive antibody, -3.338277 if two positive antibodies, -4.502697 if three positive antibodies ) + (-0.3326669 * T1DGRS2) + (0 if black, 0.6378652 if Hispanic White, -0.2740616 if Non-Hispanic White) |
